# Generative AI-assisted Bayesian-frequentist Hybrid Inference in Single-cell RNA Sequencing Analysis for Genes Associated with Alzheimer’s Disease

**DOI:** 10.64898/2026.04.17.26351142

**Authors:** Gang Han, Ao Yuan, King David Oware, Fred Wright, Raymond J. Carroll, Matthew Lee Smith, Marcia G. Ory, Dongyan Yan, Wenjie Wang, Zhe Sun, Qile Dai, Carter Allen, Andy Dang, Yushi Liu

**Author notes:** Corresponding author Correspondence to: Gang Han, Ao Yuan, Yushi Liu, (GH), (AY), liu (YL).

## Abstract

Alzheimer’s disease genomics and other high-dimensional omics studies demand powerful statistical methods, yet Bayesian inference remains underutilized despite its advantages in small-sample settings, owing to the prohibitive cost of eliciting reliable priors across thousands or millions of parameters. We propose an AI-assisted Bayesian–frequentist hybrid inference framework that couples large language model based prior elicitation with the hybrid inference theory of Yuan (2009). ChatGPT-4o is queried via a standardized prompt to assess the strength of evidence linking each gene to a disease of interest, and the response is mapped to an informative normal prior via a standardized effect-size calibration. Parameters for covariates of secondary interest are treated as frequentist parameters, preserving efficiency and avoiding sensitivity to mis-specified priors. We derive closed-form hybrid estimators under uniform and conjugate normal priors in linear models, establish their asymptotic equivalence to the frequentist and full Bayes estimators, and show in simulations that hybrid inference using unconditional variance estimation leads to high statistical power while accurately controlling the Type I error rate. Applied to single-cell RNA sequencing data from the ROSMAP cohort for Alzheimer’s disease as an example, the framework identifies biologically coherent pathways (such as gamma-secretase pathways) previously undetected. The proposed framework offers a principled and computationally scalable approach to genome-wide Bayesian analysis, with potential for broad application across omics platforms and disease settings.

## 1 Introduction

### 1.1 Alzheimer’s disease (AD) and single-cell RNA sequencing to detect relevant genes

Given global aging and dramatic projections of increased prevalence, there is considerable interest in the prevention and treatment of Alzheimer’s Disease and Related Disorders (ADRD). According to the *2025 NIH Alzheimer’s Disease and Related Dementias Research Progress Report* (https://www.nia.nih.gov/about/2025-nih-dementia-research-progress-report), more than 7 million Americans as of 2025 were living with Alzheimer’s symptoms, with this number projected to double by 2060 (Fang et al., 2025). While personal and environmental risk factors are well known, there is growing attention to cellular mechanisms and the power of advanced analytical techniques for advancing knowledge about probabilistic associations between gene expression and the presence of AD (Livingston et al., 2020). Specifically, AD has been found to be a heterogeneous neurodegenerative disorder influenced by multiple genetic and environmental factors, with the apolipoprotein E (APOE) *ϵ*4 allele representing the strongest common genetic risk factor (Safieh et al., 2019). The advent of single-cell RNA sequencing (scRNA-seq) has enabled high-resolution characterization of cellular diversity in the human brain (Nguyen et al., 2020). The Religious Orders Study (ROS) and Memory and Aging Project (MAP) cohorts (ROSMAP) provide deeply phenotyped postmortem brain samples, enabling investigation of AD-associated transcriptional variation across genetic and demographic factors such as age, sex, and race (Mathys et al., 2023; Sun et al., 2023).

### 1.2 The challenge of eliciting informative prior in Bayesian analysis

In bulk sequencing and scRNA-seq analysis, variation in technical factors with limited sample sizes can lead to low statistical power for detecting potentially significant genes. One possible solution to this problem is to use reasonable prior distribution in a Bayesian analysis. The prior elicitation plays a central role in Bayesian inference and offers a structured approach to incorporating expert or empirical knowledge into probabilistic modeling. In situations where empirical data are sparse or uncertain, a well-formed prior distribution can enhance the statistical models and result interpretation (O’Hagan, 2019; Kadane and Wolfson, 1998). Over the past three decades, several approaches have been developed to formalize the elicitation process, including structured expert judgment, hierarchical or empirical Bayesian models, and formalized group protocols (Hanea et al., 2018; Gosling, 2017). These approaches rely on experts in the field to translate substantial knowledge into quantitative distributions of probabilities, often through structured interviews, iterative calibration, and belief aggregation (Johnson et al., 2010; Garthwaite et al., 2005). Despite their theoretical rigor, elicitation in practice remains challenging because of two main reasons: First, analysts often face bias, overconfidence, inconsistent documentation and difficulties in replicating results across different practitioners (Colson and Cooke, 2017; Morgan, 2014); second, this process is time-consuming and difficult to scale up when a large number of parameters have to be defined, for example, in genomic studies with high-dimensional data (Hanea et al., 2018). Thus even if some structured guidelines are followed, the challenge of obtaining coherent and robust priors that accurately capture the domain knowledge remains a persistent obstacle to applied Bayesian analysis.

### 1.3 The promise of Artificial intelligence (AI) in eliciting prior for the Bayesian analysis of scRNA-seq data

Artificial intelligence, particularly deep learning, has become a transformative approach for analyzing large-scale omics datasets. By leveraging architectures such as convolutional neural networks, autoencoders, variational autoencoders, graph neural networks, and transformers, deep learning models can capture complex nonlinear relationships in high-dimensional molecular data, identify patterns, and predict biological outcomes that are challenging for traditional statistical approaches (Ahmed et al., 2024; Gao et al., 2022; Cui et al., 2024; Ballard et al., 2024; Marouf et al., 2025). With these advances in deep learning as well as large language models (LLMs), new opportunities are becoming available for tackling the persistent challenge of prior elicitation in the analysis of scRNA-seq data. The LLMs trained on scientific literature can synthesize dispersed domain knowledge, extract empirical evidence, and generate structured judgments similar to those of expert reasoning (Luo et al., 2025). Furthermore, generative models can suggest reasonable prior distributions and hyper-parameters consistent with prior literature, which suggests that reproducible and scalable priors can be constructed in a principled manner (Gouk and Gao, 2024). With programmatic querying and standardized prompts, such models can minimize subjectivity while increasing transparency and process the extensive biomedical literature more efficiently.

In this study, we leverage ChatGPT-4o API in a Python workflow to assess the robustness of evidence at the gene level. Specifically, we develop an algorithm to translate the generative AI’s assessment of the association between each gene and AD presence into a prior distribution for the regression parameter on the case/control status (i.e., AD vs controls) in the linear model where the gene’s expression is the outcome.

### 1.4 Implementing the hybrid Bayesian inference with covariates

The associations between multiple covariates including age, sex, APOE and gene expression are unclear, and reliable priors for these covariates are unavailable. In the statistical model, only part of the parameters have an informative prior. After eliciting the prior of case/control status for each gene, we adopt the hybrid Bayesian inference framework (Yuan, 2009) to include both frequentist and Bayesian parameters in a multivariable linear model, and make inference about the gene’s association with the presence of AD.

### 1.5 Research goal and section layout

By integrating the generative AI-based prior elicitation and Bayesian-frequentist hybrid inference, in this paper we develop an AI-assisted hybrid Bayesian inference framework to analyze scRNA-seq data taking into account informative priors and adjusting covariates, and we apply the proposed method to an AD dataset from the ROSMAP cohorts. In Section 2, we derive the key theoretical properties of the hybrid Bayesian inference in a linear model, which facilitates the implementation of the hybrid inference with AI prior elicitation presented in Section 3. The simulation results in Section 4 show that the hybrid inference is accurate, efficient and maintaining the desired Type I error rate and power. In Section 5 we analyze the ROSMAP data. Final remarks are given in Section 6.

## 2 Method

### 2.1 Hybrid Bayesian inference framework

In Statistics frequentist and Bayesian inferences are asymptotically equivalent for regular models, but for finite sample size their behaviors are different. Both types of methods have their own advantages and disadvantages. Roughly, when there is sound prior information, the Bayesian inference can have better small sample properties than the frequentist inference; when such information is lacking, the frequentist inference is better in that an incorrect informative prior can introduce bias while a non-informative prior can introduce additional uncertainty especially for small sample sizes (Han et al., 2023). In practice, often we have the situation in which, for some of the parameters there is good prior information, while for the other parameters, no such prior information exists. In this situation using the pure frequentist or full Bayesian inference is not beneficial. Previous studies have demonstrated this issue in various applications including genetic variants identification (Li et al., 2014; Yuan et al., 2014), HIV viral load modeling (Han et al., 2013, 2018), biomedical mechanic engineering design (Han et al., 2023), and scRNA-seq analyses (Han et al., 2024). To combine both methods in this situation, Yuan (2009) developed the following hybrid Bayesian inference framework, which performs a joint Bayesian-frequentist inference: a Bayesian estimate for the parameter with sound prior information and a frequentist estimate for the other parameters.

Specifically, let ***θ*** ∈ **Θ** ⊂ *R*^*p*^ (*p* > 1) denote the model parameters and *f*(*·*|***θ***) a probability density function *(pdf)* with parameters ***θ*** consisting of 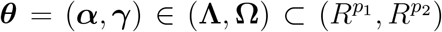, with *p* = *p*_1_ +*p*_2_, where ***α*** are *p*_1_ Bayesian parameters having prior information, and ***γ*** are *p*_2_ frequentist parameters. Let ***y***^*n*^ denote the observed data. Let 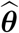 be the maximum likelihood estimate (MLE), 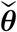 the Bayesian estimate with a prior *π*(***θ***), and 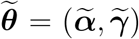 be the hybrid estimate with a prior *π*(***α***). In the hybrid inference, let 𝒟 denote the decision space for estimating ***α***, and **d** = **d**(**y**^*n*^) ∈ 𝒟 is a decision rule for ***α***. We let *L*(**d**(**y**^*n*^), ***α***) denote a loss function for estimating ***α*** by **d**(**y**^*n*^).

Using the aforementioned notations, we write the likelihood function of **y**^*n*^ as 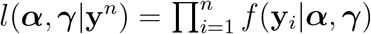 . The frequentist estimator (MLE) can be written as 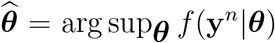 . The posterior density of ***α*** given the data and ***γ*** satisfies *π*(***α***|**y**^*n*^, ***γ***) ∝ *f*(**y**^*n*^|***α, γ***)*π*(***α***). Hybrid estimation of the frequentist parameters ***γ***, on the other hand, is based on MLE. The hybrid estimator 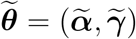 can be written as

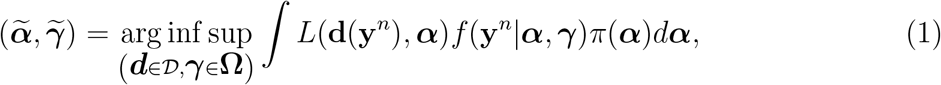

where 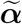 minimizes the posterior risk and 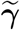 maximizes the likelihood. Yuan (2009) proved that under general conditions, the MLE, Bayesian and hybrid estimators are first order equivalent, and they are asymptotically efficient.

### 2.2 Hybrid Bayesian inference in linear models

In a linear regression model, we write the observed data with sample size *n* as (***y, X***), where ***y*** = (*y*_1_, *y*_2_, …, *y*_*n*_)^⊤^ is an *n ×* 1 vector of response values, and ***X*** is the *n × d* design matrix with (*i, j*)-th element *x*_*ij*_. We let ***x***_*i*_ denote the *d ×* 1 vector (*x*_*i*1_, *x*_*i*2_, …, *x*_*id*_)^⊤^, with *i* ∈ {1, …, *n*}. Then 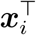 is the *i*-th row of ***X***. We let ***X***_*j*_ = (*x*_1*j*_, *x*_2*j*_, …, *x*_*nj*_)^⊤^, be the *n ×* 1 vector for all *j* = {1, …, *d}*. So 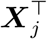 is the *j*-th column of ***X***. The linear regression parameters ***β*** = (*β*_1_, *β*_2_, …, *β*_*d*_)^⊤^ can also be written as 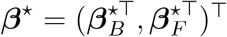 where ***β***_*B*_ and ***β***_*F*_ are Bayesian and frequentist parameters in the hybrid inference, respectively. We let ***X***_*B*_ and ***X***_*F*_ denote the columns of ***X*** corresponding to ***β***_*B*_ and ***β***_*F*_, respectively. In the linear regression model, ***y*** is a realization of a random vector ***Y*** modeled as

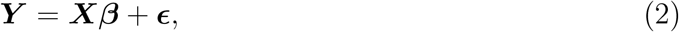

where elements in ***ϵ*** = (*ϵ*_1_, …., *ϵ*_*n*_)^⊤^ are independent and identically distributed (*i*.*i*.*d*.) Normal (0, *σ*^2^). In the estimation of ***β***, we let 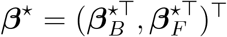 denote the true or ideal value. Similar to the estimators of ***θ*** defined in section 2.1, we let 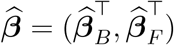 denote frequentist least squares estimate (LSE), 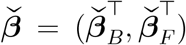 the Bayesian estimate, and 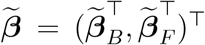 the hybrid Bayesian estimate. In Bayesian inference the prior of all the parameters can be written as *π*(***β***, *σ*^2^). In hybrid inference the prior can be written as *π*(***β***_*B*_) assuming *σ*^2^ is a frequentist parameter. In case ***β*** has a uniform prior, we let ***a*** and ***b*** denote the lower bound and upper bounds, where ***a*** = (*a*_1_, *a*_2_, …, *a*_*d*_), ***b*** = (*b*_1_, *b*_2_, …, *b*_*d*_) and *π*(***β***) = *U*[***a, b***]. If ***β*** has a normal conjugate prior, we denote the prior distribution in Bayesian inference as *π*(***β***) ∼ *N*(***µ*, Σ**) and in hybrid inference as *π*(***β***_*B*_) ∼ *N*(***µ***_*B*_, **Σ**_*B*_). Moreover, we let *ϕ*(*·*) and Φ(*·*) denote the *pdf* and cumulative density function of the standard normal distribution, respectively. We let 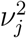 denote the (*j, j*)-th element of the *d × d* matrix (***X***^⊤^***X***)^−1^; note that 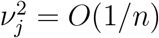 for all *j* = {1, …, *d*} and 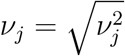.

Using the above notations, Theorem 1 below specifies the estimation from frequentist, Bayesian, and hybrid inferences with three different loss functions (i.e., 0-1 loss, absolute error loss, and squared error loss) and a uniform prior on ***β*** under the linear model (2), while Theorem 2 specifies the estimation with a normal conjugate prior on ***β***. Proofs of the two theorems are given in the Appendix.

#### Theorem 1.

*Assume in the regression model (2)*, ***β*** *has a uniform prior π*(***β***) ∼ *U*[***a, b***], *and* [***a, b***] *covers* ***β***^⋆^.

1. *Under a 0-1 loss function*,

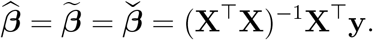
2. *Under the absolute error loss, the component-wise Bayesian estimator is*

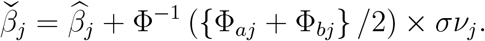
3. *Under the squared error loss, the component-wise Bayesian estimator is*

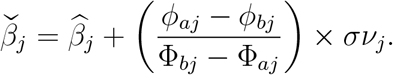

It is worthwhile to note that in Theorem 1, with *v*_*j*_ = *O*(*n*^−1*/*2^), if 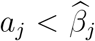 and 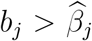, then 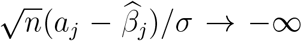, and 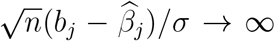 with *n* → ∞. So Φ_*bj*_ − Φ_*aj*_ → 1, *ϕ*_*bj*_ − *ϕ*_*aj*_ → 0, and 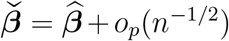. As a result, 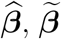 and 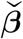 are asymptotically equivalent.

According to the Bernstein-von Mises theorem (Doob, 1949), under general conditions 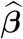 and 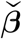 are asymptotically equivalent. One of the conditions in the Bernstein-von Mises theorem requires that the support of the model does not depend on the parameter. In Theorem 1, however, the support (*a*_*j*_, *b*_*j*_) does depend on the parameter *β*_*j*_, for all *j* ∈ {1, …, *d*}. The three estimators are still asymptotically equivalent.

#### Theorem 2.

*Assume in the regression model (2), in the Bayesian inference* ***β*** *has a conjugate prior π*(***β***) ∼ *N*(***µ*, Σ**). *In the hybrid inference* ***β***_*B*_ *has a conjugate prior π*(***β***_*B*_) ∼ *N*(***µ***_*B*_, **Σ**_*B*_). *Then with the 0-1 loss, absolute error loss, and squared error loss, the Bayesian estimate can be written as*

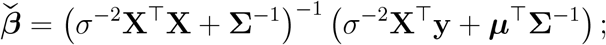

*and the hybrid estimate can be written as*

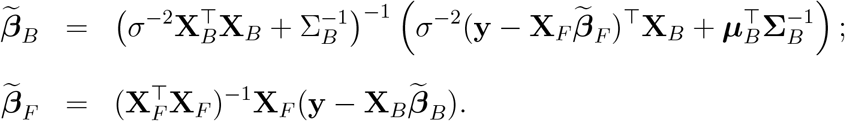

With the above two theorems, we further discuss a special condition under which the hybrid inference and Bayesian inference lead to an identical estimate of the regression parameters in the following corollary.

#### Corollary 1.

*Given the linear model (2) and with squared error loss, the hybrid inference and full Bayesian inference have the identical point estimate of* ***β*** *for any n, d*, ***µ***_*B*_, **Σ**_*B*_ *if the prior of* ***β***_*F*_ *is constant with no boundary in the full Bayesian inference, which corresponds to* ***a*** → **−∞** *and* ***b*** → **∞** *for the uniform prior π*(***β***) = *U*[***a, b***] *in Theorem 1, or* diag(**Σ**_*F*_) → **∞** *for the conjugate prior π*(***β***_*F*_) ∼ *N*(***µ***_*F*_, **Σ**_*F*_) *in Theorem 2*. ■

As to be shown in Section 3, this corollary can greatly reduce the computation time of the hybrid inference if the conditions are met. Proofs of Theorems 1, 2, and Corollary 1 are given in Appendix A.

### 2.3 A practical example with four independent variables and an informative prior on one regression parameter

We next discuss the hybrid inference in a practical example with four independent variables, and derive the asymptotic and finite sample variances of 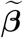, assuming *σ*^2^ is treated as a frequentist parameter. In a linear model for a gene expression, we assume there are four independent variables including 1) case/control status, 2) APOE, 3) age, and 4) sex. In this example, ***β***_*B*_ = *β*_1_, ***β***_*F*_ = (*β*_0_, *β*_2_, *β*_3_, *β*_4_)^⊤^, ***X***_*B*_ = ***X***_1_, and ***X***_*F*_ = [***X***_0_, ***X***_2_, ***X***_3_, ***X***_4_], where *β*_0_ is the intercept and ***X***_0_ is an *n ×* 1 vector of 1’s. The true value ***β***^⋆^ can be written as 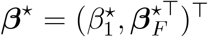. Given ***β***_*F*_, we let ***z*** denote the frequentist predictor without the Bayesian parameters, i.e., ***z*** = ***X***_*F*_ ***β***_*F*_ = (*z*_1_, *z*_2_, …, *z*_*n*_)^⊤^, where *z*_*i*_ = *β*_0_ + *β*_2_*x*_*i*2_ + *β*_3_*x*_*i*3_ + *β*_4_*x*_*i*4_, for all *i* ∈ {1, 2, …, *n*}. Similarly we let 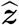 and 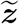 denote the frequentist predictor and hybrid predictor without the Bayesian parameters, i.e., 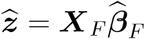 and 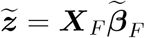 . We use *I*(*·*) to denote an indicator function. Finally we let 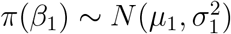 denote the prior on *β*_1_.

With the above notations, we derive the posterior distribution of *β*_1_ given parameters (***β***_*F*_, *σ*^2^), data (***X, y***), and the two priors in Theorems 1&2. In model (2) and for each *i*, we have 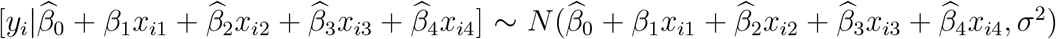, for all *i* = 1, …, *n*. We assume the prior *π*(*β*_1_) ∼ Uniform[*a*_1_, *b*_1_], (*a*_1_, *b*_1_) → (−∞, ∞). The posterior density for *β*_1_ can be derived as

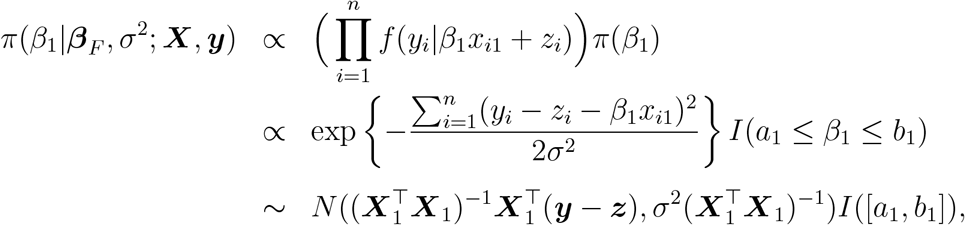

which is the truncated normal distribution on [*a*_1_, *b*_1_].

Assume the prior is the normal conjugate prior in Theorem 2, *π*(***β***) ∼ *N*(***µ*, Σ**). In the Bayesian inference, according to literature the posterior distribution of ***β*** is

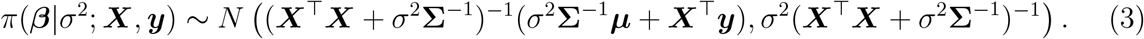

In the hybrid inference, the posterior of *β*_1_ can be written as

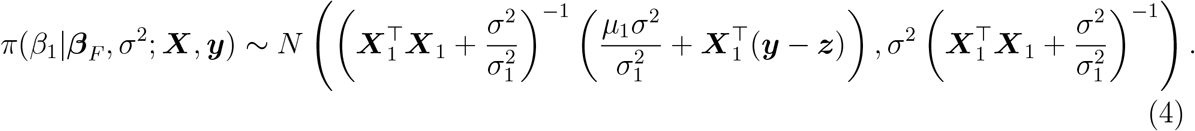

According to Theorem 2 and the posterior distribution in (4), the point estimates and confidence intervals of *β*_1_ from Bayesian and hybrid inference can be obtained using the posterior means and variances. It is worthwhile to note that in the hybrid inference, this confidence interval is conditional on the frequentist parameter estimates 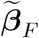.

### 2.4 The conditional variance 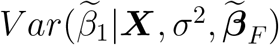 and the unconditional variance 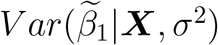

To construct the unconditional confidence interval of *β*_1_ without conditioning on 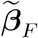, we next investigate the difference between conditional and unconditional variances, and derive the unconditional estimation variances of *β*_1_, including 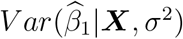 and 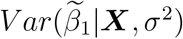 assuming an informative conjugate normal prior. We start from the asymptotic variance and then move to the finite sample variance.

Regarding the asymptotic variance, in the frequentist inference we can re-arrange the MLEs’ order as 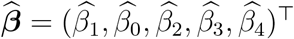, and make the same re-arrangement in ***X***, i.e., having the observations of ***X***_1_ in the first column, followed by a column of ones and observations from the covariates ***X***_2_, ***X***_3_, and ***X***_4_. Then the variance matrix of 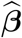 can be defined as a matrix with elements (*ω*^11^, **Ω**^12^, **Ω**^12^, **Ω**^22^). According to Yuan (2009), given the priors in Theorem 2, the LSE 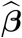, the full Bayesian estimator 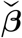, and the hybrid estimator 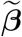 are asymptotically equivalent as *n* → ∞. The asymptotic estimation variance can be written as

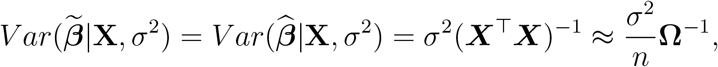

where

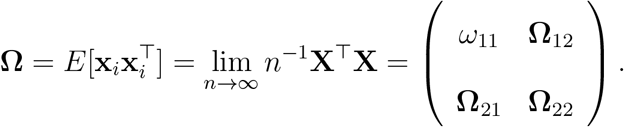

Denote

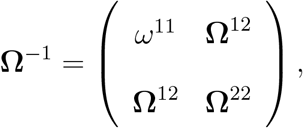

then *ω*_*11*_ is the variance of 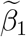 . Then 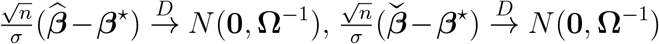, and 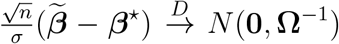 . Based on the covariance matrix 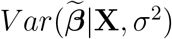, the asymptotic variance of 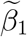 is thus 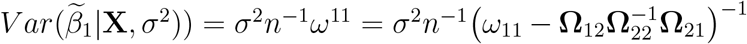. But conditioning on frequentist parameter estimates 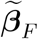, the asymptotic conditional variance of 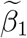 is 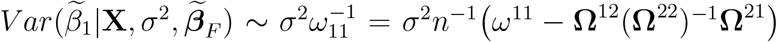. Note that 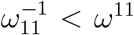 because 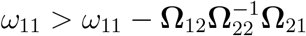 or 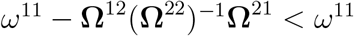. Difference in the asymptotic conditional and unconditional variance can be quantified as 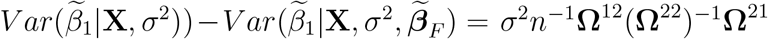, and we can see the previously derived posterior distribution (4) has the conditional variance.

For the finite sample variance, the estimation variance of the LSE 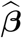 remains 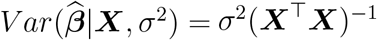. In Theorem 2, 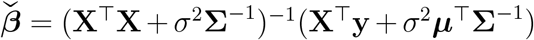. Then the estimation variance from the Bayesian inference 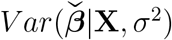 can be derived as

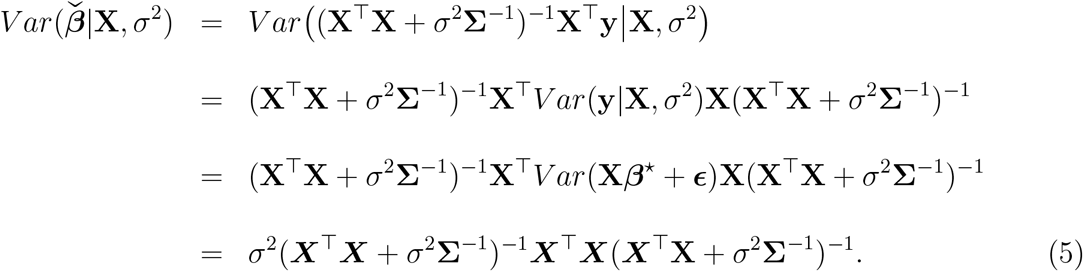

Note that (5) can be approximated by *σ*^2^*n*^−1^**Ω**^−1^ with a large sample size as in the afore-mentioned asymptotic inference, because (5) can be written as *σ*^2^(**X**^⊤^**X** +*σ*^2^**Σ**^−1^)^−1^(**X**^⊤^**X** + *σ*^2^**Σ**^−1^ −*σ*^2^**Σ**^−1^)(**X**^⊤^**X**+*σ*^2^**Σ**^−1^)^−1^ = *σ*^2^(**X**^⊤^**X**+*σ*^2^**Σ**^−1^)^−1^ −*σ*^4^(**X**^⊤^**X**+*σ*^2^**Σ**^−1^)^−1^**Σ**^−1^(**X**^⊤^**X**+ *σ*^2^**Σ**^−1^)^−1^ ≈ *σ*^2^*n*^−1^**Ω**^−1^ +*O*(*n*^−2^). Then 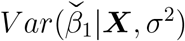 is the corresponding diagonal element of 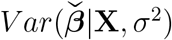.

Next we derive the finite sample variance of the hybrid estimator. By Theorem 2 and the posterior distribution (4), 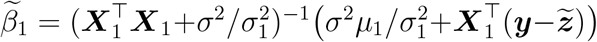, where 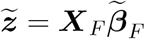. If 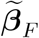 and 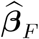 are close, 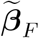 can be written as 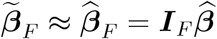, where ***I***_*F*_ is the (d − 1) *× d* matrix with the row corresponding to *β*_1_ removed. Then

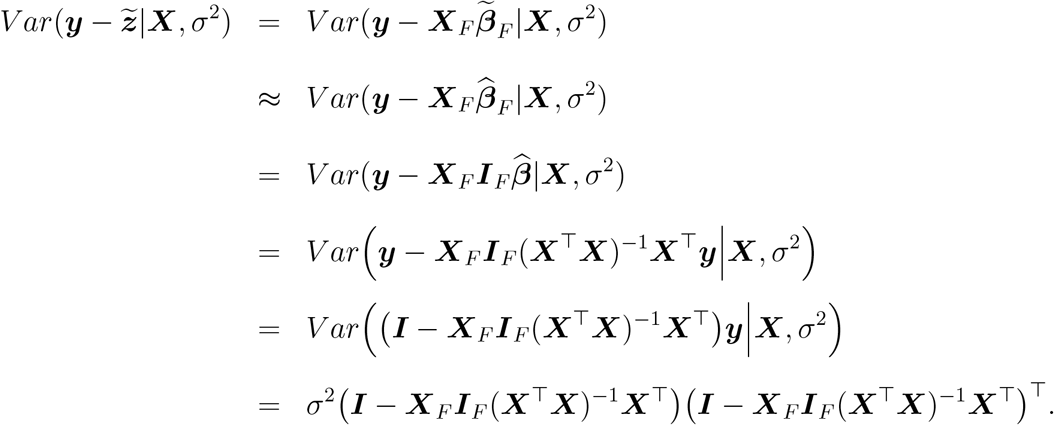

The unconditional variance of 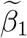 can be derived as

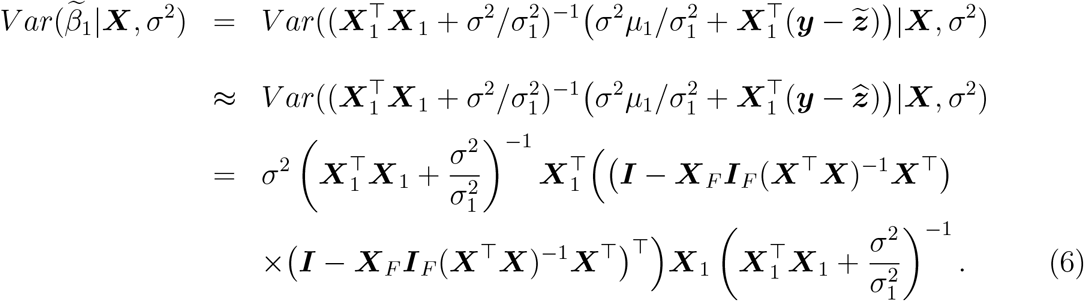

Alternatively, numerical methods with a large number of simulation iterations (e.g., bootstrap methods) could be utilized to evaluate the estimation variance of 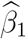 and 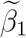. Last but not least, the estimation of the *σ*^2^ in the hybrid inference is based on the least square estimate, i.e. 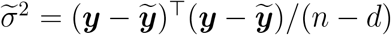, where 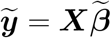and *d* = 5.

## 3 Implementation

### 3.1 AI tools for eliciting priors

After collecting the ROSMAP data of patient demographics and genes linked to AD (Mathys et al., 2023; Sun et al., 2023), we developed a standardized prompt for ChatGPT-4o to collect evidence from previous genetic, genomic, and proteomic studies. We used generative AI tools to gauge the association between genes and AD by categorizing the association level as a five-point ordinal scale: 1 = no evidence, 2 = weak, 3 = moderate, 4 = moderate-to-strong, 5 = strong. A Python program was implemented utilizing the *OpenAI* Application Programming Interface (API) by 1) querying every gene iteratively, 2) parsing the AI’s response, and 3) deriving the association level. The association levels of all the genes were saved as a numerical vector. This automated process provided a consistent method for searching available information about each gene and transforming textual records into quantitative priors. The resulting evidence level vector was then used as structured informative priors in our Bayesian–frequentist hybrid analysis. Given the significance and challenges of obtaining dependable prior distributions in Bayesian analyses (Johnson et al., 2010), this novel use of the generative AI could be useful to run regression analysis with informative priors (Riegler et al., 2025) and to advance the application of AI-based genetic and genomic studies in AD research, according to Mishra and Li (2020). The OpenAI API query implemented in Python is provided in Appendix B.

### 3.2 Computation steps

Our proposed steps below can incorporate the AI inputs with the hybrid Bayesian inference computation proposed in Han et al. (2023).

#### Step 1

Implement the generative AI to gauge the association between the gene and AD from existing evidence in five levels. Levels 1-5 correspond to none, weak, moderate, moderate-to-strong, and strong, respectively.

#### Step 2

Calculate the standard deviation in this gene’s expression from a prior dataset for the control group and case group. Let *S*^2^ denote the pooled variance. Use the prior data to decide whether AD case is on average having higher or lower expression than control. Let *I*_*c*_ = 1 if the gene expression in the case group is higher than the control group, and *I*_*c*_ = −1 if the gene expression in the control group is higher than the case group.

#### Step 3

Construct the prior for *β*_1_ using the mean difference of the genes between the AD and control in terms of Cohen’s d, which are chosen based on existing literature (Cohen, 2013; Kraemer and Kupfer, 2006; Sawilowsky, 2009). The priors of *β*_1_ at different Cohen’s d values are set to:

- Cohen’s d = 0 for level 1, *π*(*β*_1_) ∼ *N*(0, *S*^2^);
- Cohen’s d = 0.2 for level 2, *π*(*β*_1_) ∼ *N*(0.2 *× I*_*c*_ *× S, S*^2^);
- Cohen’s d = 0.5 for level 3, *π*(*β*_1_) ∼ *N*(0.5 *× I*_*c*_ *× S, S*^2^);
- Cohen’s d = 0.9 for level 4, *π*(*β*_1_) ∼ *N*(0.9 *× I*_*c*_ *× S, S*^2^);
- Cohen’s d = 1.5 for level 5, *π*(*β*_1_) ∼ *N*(1.5 *× I*_*c*_ *× S, S*^2^).
- Cohen’s d = 3.25 for level 6, *π*(*β*_1_) ∼ *N*(3.25 *× I*_*c*_ *× S, S*^2^).

#### Step 4

With the above conjugate prior in the form of 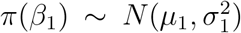, run the hybrid Bayesian inference with the procedure specified in Han et al. (2023), where the expectation-maximization algorithm proposed in Han et al. (2023) can be simplified by Corollary 1. The initial estimate of ***β***_*F*_ = (*β*_0_, *β*_2_, *β*_3_, *β*_4_) and *σ*^2^ can be the LSE, denoted as ***β***_*F* (0)_ and 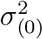 . At the *k*-th iteration, the estimates of *β*_1_, ***β***_*F*_ and *σ*^2^ can be updated as *β*_1(*k*)_, ***β***_*F* (*k*)_ and 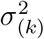 using the following three equations

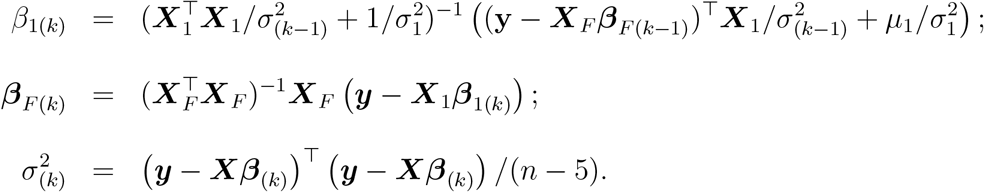

According to Corollary 1, the computation for point estimation in **Step 4** can be further simplified assuming a non-informative prior on *β*_0_, *β*_2_, *β*_3_, and *β*_4_, if the conditions in Corollary 1 are met. The posterior distribution of ***β*** is the Gaussian distribution in (3), leading to the point estimate 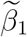 with 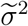 approximated by 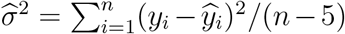. The conditional variances of 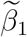 can be calculated using 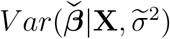 in equation (5), while the unconditional variance 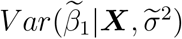 can be calculated using equation (6).

## 4 Simulation examples

### 4.1 Simulation settings

In this section we simulate the data from a linear model and then run the frequentist, Bayesian, and hybrid inference to evaluate the point estimation and estimation variance. We further compare the variance from the point estimates in simulation and the estimation variance to understand if the variability was estimated with any bias, where an overestimation of the variance leads to overly conservative test of the regression parameter and less power, while an underestimation of the variance leads to high power but Type I error inflation.

Following (2), the response, *y*_*i*_, is generated from a normal distribution with mean *β*_0_ + *β*_1_*x*_*i*,1_ + *β*_2_*x*_*i*,2_ + *β*_3_*x*_*i*,3_ + *β*_4_*x*_*i*,4_ and variance *σ*^2^. The true values of the regression parameters are set to ***β*** = (*β*_0_, *β*_1_, *β*_2_, *β*_3_, *β*_4_)^⊤^ = (7, 0.5, 0, 0, 0)^⊤^. The standard deviation *σ* is set to 0.58 with an effect size (two sample Cohen’s d) for *β*_1_ equal to 0.86 = *β*_1_*/σ* = 0.5/0.58, which corresponds to a moderate-to-strong effect size as described at level 4 of Step 3 in Section 3.2. With a sample size *N*, we let half of the *x*_*i*,1_ be 0 and the other half be 1. Regarding the covariates, we generate (*x*_*i*,2_, *x*_*i*,3_, *x*_*i*,4_) with the distributions *x*_*i*,2_ ∼ *N*(70, 30^2^), *x*_*i*,3_ ∼ *Bernoulli*(0.5) and *x*_*i*,4_ ∼ *Bernoulli*(0.5). With (*β*_2_, *β*_3_, *β*_4_) = (0, 0, 0), the three covariates have no impact on the outcome *y*_*i*_. In the Bayesian and hybrid inference, the prior of *β*_1_ follows a normal distribution with mean *µ*_1_ and variance 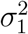. The regression parameters (*β*_0_, *β*_2_, *β*_3_, *β*_4_) have non-informative prior in the Bayesian inference and are set to frequentist parameters in the hybrid inference. Finally we used the parametric empirical Bayes approach (Morris, 1983) to set the value of *σ*^2^ in Bayesian inference.

In Tables 1 and 2, values of the prior’s mean *µ*_1_ are set to 0, 0.5, and 1. The values of the prior’s standard deviation *σ*_1_ are set to 0.1, 0.7, and 1, corresponding to strong, moderate and weak prior information, respectively. The sample size *N* is set to 50 and the number of simulations is 10,000. We summarize the point estimate with standard deviation (SD) from simulation in Table 1 and the Standard Error Estimate (SEE) with interquartile range (IQR) in Table 2. Here the estimation variance from the hybrid inference is from the unconditional variance in equation (6).

**Table 1:** Point estimate. Mean (SD) from 10K iterations; *σ* = 0.58, *N* = 50 in each iteration. True value in the simulation *β*_1_ = 0.5. The prior distribution of *β*_1_ is 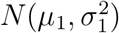; ▲ indicates the prior mean is equivalent to the true parameter value.

| | $\mu_1 = 0$ | $\mu_1 = 0.5 \blacktriangle$ | $\mu_1 = 1$ |
| --- | --- | --- | --- |
| Frequentist | 0.499 (0.169) | 0.499 (0.169) | 0.502 (0.170) |
| Bayesian and Hybrid, $\sigma_1 = 1$ | 0.485 (0.164) | 0.502 (0.164) | 0.516 (0.165) |
| Bayesian and Hybrid, $\sigma_1 = 0.7$ | 0.468 (0.161) | 0.499 (0.160) | 0.525 (0.161) |
| Bayesian and Hybrid, $\sigma_1 = 0.1$ | 0.132 (0.051) | 0.502 (0.045) | 0.867 (0.050) |

**Table 2:** Estimation variance. Median [Interquartile range] from 10K iterations; *σ* = 0.58, *N* = 50 in each iteration. True value in the simulation *β*_1_ = 0.5. The prior distribution of *β*_1_ is 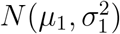; ▲ indicates the prior mean is equivalent to the true parameter value.

| | $\mu_1 = 0$ | $\mu_1 = 0.5 \blacktriangle$ | $\mu_1 = 1$ |
| --- | --- | --- | --- |
| Frequentist | 0.168 [0.157, 0.181] | 0.168 [0.156, 0.179] | 0.168 [0.157, 0.181] |
| Bayesian and hybrid, $\sigma_1 = 1$ | 0.166 [0.155, 0.178] | 0.165 [0.154, 0.176] | 0.166 [0.155, 0.179] |
| Bayesian and hybrid, $\sigma_1 = 0.7$ | 0.163 [0.152, 0.176] | 0.163 [0.151, 0.175] | 0.164 [0.152, 0.175] |
| Bayesian, $\sigma_1 = 0.1$ | 0.086 [0.084, 0.088] | 0.086 [0.084, 0.088] | 0.086 [0.084, 0.088] |
| Hybrid, $\sigma_1 = 0.1$ | 0.072 [0.071, 0.073] | 0.072 [0.071, 0.073] | 0.072 [0.071, 0.073] |

### 4.2 Point estimation

According to Table 1, the frequentist inference is invariant with prior settings that have a significant impact on Bayesian and hybrid inferences. Consistent with Corollary 1, Bayesian and hybrid inference point estimates are identical in all the simulation runs. If the prior mean is equivalent to the true value of *β*_1_, i.e. *µ*_1_ = *β*_1_ = 0.5, all estimators are unbiased. The Bayesian and hybrid estimators, however, are more efficient with less variability in the estimation than the frequentist estimator: The frequentist estimator had standard deviation from the simulation being 0.169, and 0.169 > 0.164 > 0.160 > 0.045, where 0.164, 0.160, and 0.045 were the standard deviations from hybrid (and Bayesian) inference, for *σ*_1_ = 1, *σ*_1_ = 0.7, and *σ*_1_ = 0.1, respectively, at *µ*_1_ = 0.5. If the prior mean is *incorrectly* specified as *µ*_1_ = 0 or *µ*1 = 1, Bayesian and hybrid inference could be biased, with lower prior variance 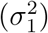 corresponding to more severe bias. The standard deviation from the simulation runs decreases with a smaller prior variance for all values of the prior mean *µ*_1_. For example, with *µ*_1_ = 1, the standard deviations were 0.050, 0.161, and 0.165 for *σ*_1_ = 0.1, 0.7, and 1, respectively. This finding confirms that a stronger prior information (or smaller *σ*_1_) leads to a more stabilized point estimate of *β*_1_.

### 4.3 Estimation variance

In Table 2, the estimation variance did not depend on the prior mean *µ*_1_ in Bayesian and hybrid inferences, which is consistent with the derivation in Section 2. The Bayesian and hybrid estimation variances were less than that from the frequentist inference in all the simulation runs, which was due to the prior information. When the prior variance 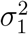 was relatively large with *σ*_1_ = 1 or *σ*_1_ = 0.7, the Bayesian and hybrid estimation variance were equivalent. However, with a strong prior (the last two rows of Table 2 where *sigma*_1_ = 0.1), the hybrid inference had a smaller estimation variance than the Bayesian inference (standard error 0.072 < 0.086). This finding is consistent with the existing literature (Han et al., 2023) that the Bayesian inference could introduce additional variability compared with the hybrid inference, given that frequentist parameters all had non-informative priors.

### 4.4 Type I error rate and power

We further discuss the Type I error rate and power using Tables 1 and 2. In Table 1, the SD indicates the true variability of the point estimate. A standard error estimate from Table 2 close to the SD in Table 1 indicates that the variance estimation and a corresponding confidence interval is generally accurate. If a standard error estimate in Table 2 is substantially higher than the corresponding SD in Table 1, it indicates overly conservative inference for *β*_1_ with low statistical power. Otherwise, if the standard error estimate is less than the SD in Table 1, the inference inflates the Type I error rate. Comparing the estimated SEE in Table 2 and the SD in Table 1, the frequentist inference, Bayesian and hybrid inference with *σ*_1_ = 1 and *σ*_1_ = 0.7 all had their SEE comparable to the corresponding SD. For example, at *µ*_1_ = 0.5, the SEE and SD were 0.168 and 0.169 respectively from the frequentist inference, and the SEE and SD were 0.165 and 0.164 from the Bayesian and hybrid inference at *σ*_1_ = 1. However, with a strong prior where *σ*_1_ = 0.1, the SEE from the hybrid inference (i.e., 0.072) is less than the SEE from Bayesian inference (i.e., 0.086) for all the values of *µ*_1_ in Table 2. And the SEE from the hybrid inference (0.072) was closer to (but never less than) the corresponding SD values in Table 1, which were 0.045 to 0.051. This indicates that the hybrid inference with unconditional variance can be more powerful than the Bayesian inference without compromising the Type I error rate, when the prior provides strong information.

### 4.5 Additional simulations available in supplementary files

In the supplementary files we presented the simulation results under more scenarios, including the sample size *N* set to 50, 100 and 300, higher variance in the response *σ* = 3, and the SEE was also reported using the conditional variance from the hybrid inference. Moreover, the true value of *β*_1_ was set to 0, 0.5, and 1, and *µ*_1_ was set to 0, 0.2, 0.5, and 1 in the simulation. These additional results are consistent with the findings in Tables 1 and 2. Specifically, with less information from data (lower N, higher *σ*^2^), the impact of the prior 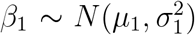 is higher on the point estimate, especially with a strong informative prior. The SEE from the conditional variance of the hybrid inference is generally less than that from the unconditional variance. The SEE from the conditional variance, however, could be less than the corresponding SD of the estimates from the simulation, indicating that the hybrid inference with conditional variance may suffer from Type I error inflation as a cost of its higher statistical power. On the other hand, the hybrid inference with unconditional variance is free from the Type I error inflation issues.

## 5 Application

In this section, we apply the Bayesian–frequentist hybrid inference to inhibitory neurons single-nucleus RNA sequencing (snRNA-seq) data from the ROSMAP database introduced in Section 1. The s**n**RNA-seq is a complementary technique to s**c**RNA-seq, adapted for frozen and postmortem tissue, enabling transcriptomic profiling of cell types that are difficult to dissociate intact.

### 5.1 Data source and cell-level quality control preprocessing

The snRNA-seq data were obtained from the public domain. The ROSMAP snRNA-seq dataset was generated from 427 postmortem human prefrontal cortex samples by Mathys and colleagues (Mathys et al., 2023). We also used another snRNA-seq dataset, from 21 prefrontal cortex tissue samples by Lau and colleagues (Lau et al., 2020), to generate the variability for prior construction of each gene. For both datasets, we used Braak stage, a severity measure of neurofibrillary tangle pathology in AD (Braak and Braak, 1991), to dichotomize samples into “mild/none” (Braak stages 0-2) and “severe AD” (Braak stages 4-6) groups. Both datasets were processed using Seurat R package for quality control (QC) metric calculation and pseudo-bulk aggregation (Stuart et al., 2019; Hao et al., 2024). Pseudo-bulk samples were then normalized using *EdgeR* R package (Chen et al., 2025) and limma-voom R packages (Ritchie et al., 2015).

Cell-level QC metrics were calculated including the percentage of reads mapping to mitochondrial genes, MALAT1 expressions, long non-coding RNAs, and ribosomal protein genes. Principal component analysis was performed on these cell-level QC features, and the first three principal components were aggregated to the patient level by taking the mean PC scores across all cells within each donor. For each cell type, the single-cell expression data were aggregated to pseudo-bulk samples by summing raw counts across all cells within each donor. Samples with fewer than 20 cells per cell type were excluded from downstream analysis to ensure robust pseudo-bulk estimates. Gene expression was filtered to retain only genes expressed at > 1 count per million (CPM) in at least 15% of samples. Additionally, only protein-coding genes based on Ensembl gene biotype annotation were retained for normalization. Library size normalization was first performed using the trimmed mean of M-values (TMM) method implemented in the EdgeR package (Chen et al., 2025). Expression data were then transformed using the voom method with sample-specific quality weights (Law et al., 2014) using the limma R package (Ritchie et al., 2015). Additionally, the standard deviation per gene for each group for Lau’s data were calculated as the potential input of variability for prior. The final processed data included: (1) ROSMAP dataset: 13,649 genes from 203 severe AD and 113 mild/none AD samples for Inhibitory neurons; (2) Data from Lau et al. (2020): 14,908 genes from 12 severe AD and 9 mild/none AD samples for Inhibitory neurons. The ROSMAP gene list was further filtered based on average count per cell greater than 0.5 to ensure we have enough expression and the filtration further resulted in 3885 genes for prior generation.

### 5.2 AI-assisted prior elicitation

With the binary case/control status and each gene’s expression data, we wrote ChatGPT prompt query in Python to interact with ChatGPT API (gpt-4o api version=2024-10-21) on the Azure platform. Each gene was iteratively included in each prompt to derive the informative prior and neuron-specific informative prior on how tight the candidate gene is associated with AD. The strength of such evidence was then categorized into 5 different levels with level 5 as the strongest evidence. The results were aggregated into a csv format for subsequent use. The prompt is listed in Appendix B.

### 5.3 Data analysis using the prior elicited by ChatGPT-4o

We conduct differential gene expression analyses for inhibitory neurons with the proposed hybrid inference. Similar to the simulation examples in Section 4, in the regression model, *y*_*i*_ is the observed gene expression, and (*x*_*i*,1_, *x*_*i*,2_, *x*_*i*,3_, *x*_*i*,4_) correspond to case/control status, age, sex, and APOE, respectively. The frequentist parameters include (*β*_0_, *β*_2_, *β*_3_, *β*_4_, *σ*^2^) and the Bayesian parameter is *β*_1_ with prior 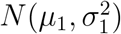.

Analysis of each gene has been conducted in four different prior settings, including

1. No evidence (frequentist inference): We assumed the prior strength as 0 (no evidence) with the variance equivalent to that of the gene in Lau’s data (Lau et al., 2020);
2. Non-informative prior: we assumed the AD associated strength from ChatGPT with a large variance (pooled variance 10^8^);
3. Informative prior: we assumed the AD associated strength from ChatGPT with the variance calculated based on Lau’s data (Lau et al., 2020);
4. Neuron-specific informative prior: we assumed the AD associated strength specific to neuron from ChatGPT with the variance based on Lau’s data.

Bayesian analysis with each prior setting was conducted using ROSMAP snRNA-seq data modeling the gene expression with dichotomized Braak stages (Mathys et al., 2023) adjusting baseline age, sex, APOE genotype (E4 carrier or not). We filtered the top ranked genes based on the absolute log fold change larger than 0.25 and false discovery rate (FDR) less than 0.2 and the gene lists were further used for pathway analysis using Metacore (Clarivate).

### 5.4 Results and interpretations

The pathways with FDR less than 0.1 were tabulated in Table 3. In the no evidence or frequentist setting (setting 1), none of the genes had FDR less than 0.2. With the non-informative prior (setting 2 in Section 5.3), no pathways were identified to be significant. As a result, the AI-assisted priors increased the sensitivity of the pathway analysis regarding the AD mechanisms. With the informative prior (setting 3 in Section 5.3), two gamma secretase related pathways were identified. Gamma-secretase cleaves amyloid precursor protein, which generates amyloid-beta (A*β*) peptides (Haapasalo and Kovacs, 2011) to form amyloid plaques, a well-known hallmark of AD (Dries and Yu, 2008). Another cleavage target for gamma secretase was Notch, and evidence showed inhibition of gamma-secretase led to changes in neurite morphology and synaptic structure via Notch signaling pathway (Figueroa et al., 2002). Moreover, the heterochromatin protein 1 (HP1) protein family was significantly associated to AD in our analysis. This is consistent with findings reported by Neuner et al. (2019) that chromatin remodeling pathway was affected by Tau protein in AD brains, and reduction in heterochromatin markers such as HP1 was also observed.

**Table 3:** Q values from the informative prior and neuron-specific informative prior in six different pathways. Pathway 1: Gamma-secretase proteolytic targets; Pathway 2: Gamma-Secretase regulation of neuronal cell development and function; Pathway 3: Neurophysiological process ACM1, ACM3 and ACM5 signaling in the brain; Pathway 4: Transcription Role of HP1 family in transcriptional silencing; Pathway 5: Neurophysiological process Dynein-dynactin motor complex in axonal transport in neurons; Pathway 6: Prolactin/JAK2 signaling in breast cancer. “⋆⋆” indicates a q-value ≤ 0.05; “⋆” indicates a q-value ∈ (0.05, 0.1).

| Different Pathways | Informative prior | Neuron-specific informative prior |
| --- | --- | --- |
| Pathway 1 | 5.75E-04** | 1.75E-03** |
| Pathway 2 | 6.81E-04** | 1.78E-03** |
| Pathway 3 | 0.04** | 0.07* |
| Pathway 4 | > 0.1 | 3.65E-03** |
| Pathway 5 | 0.10* | > 0.1 |
| Pathway 6 | 0.10* | > 0.1 |

The hybrid analysis based on the neuron specific informative prior (setting 4 in Section 5.3) yielded more interesting results. The HP1 protein pathway became more significant than the analysis based on non-informative prior. Another unique pathway close to being significant was neurofilaments in axon growth and synapses. Existing works have indicated that neurofilaments are neuron-specific intermediate filaments that play a key role in regulating axon caliber (Lee and Cleveland, 1996). In AD, abnormal distribution and hyperphosphorylation of neurofilament proteins could be linked to axonal damage, and the development of neurofibrillary tangles, another important hallmark of AD pathology (Fernandez-Martos et al., 2015; Mitew et al., 2013). Thus it is not surprising to see neuron specific pathways from our Bayesian analysis using neuron specific prior. Additional pathway analysis with q-values close to 0.1 is given in the supplementary materials. In conclusion, the proposed approach can enable robust estimation and powerful detection of AD-associated transcriptional changes while accounting for key genetic, demographic, and technical covariates, which generates cell type-specific molecular signatures of AD pathology.

## 6 Final Remarks

In this paper, we developed a statistical method to mitigate the challenges of (i) prior elicitation, and (ii) incorporating covariates without informative priors, in the analysis of scRNA-seq (and snRNA-seq) data, and this method was illustrated in the analysis of ROSMAP data to identify plausible mechanisms linking gene expression to AD outcomes. Specifically, we proposed an AI-assisted hybrid Bayesian inference framework that integrates two major methodological components, generative AI-based prior elicitation and Bayesian-frequentist hybrid inference, which complement each other well in our analysis presented in Section 5. Regarding the prior elicitation, expert elicitation is prohibitively time-consuming and difficult to scale in high-dimensional genomic data analysis, but the proposed AI-assisted prior elicitation leverages ChatGPT-4o to synthesize dispersed evidence from the biomedical literature and translate it into structured priors. On the other hand, the hybrid Bayesian inference module, building on the theoretical foundation of Yuan (2009) and the computational framework of Han et al. (2023), incorporates these AI-generated priors for the parameter of primary scientific interest while treating nuisance covariates as frequentist parameters. This hybrid structure preserves the efficiency gains of Bayesian inference and avoids the well-known sensitivity of full Bayesian analysis to mis-specified priors on parameters for which no reliable prior information exists, and the hybrid inference is more efficient than the full Bayesian analysis using non-informative prior, according to the simulation in Section 4 and Han et al. (2023). To our knowledge, the derivation and implementation of the unconditional variance in Equation (6) are novel contributions that have not been previously reported in the literature. Together, the two components form a coherent and practically deployable pipeline for genome-wide Bayesian analysis.

Our simulation results confirm that the hybrid inference framework achieves controlled Type I error rates and improved statistical power relative to both frequentist and full Bayesian approaches when the prior information is accurate or approximately correct. Consistent with Theorems 1 and 2, the three estimators are asymptotically equivalent, but meaningful finite-sample efficiency gains are achievable with informative priors, particularly in the small-to-moderate sample size settings typical of single-cell studies. The use of unconditional rather than conditional variance for inference, as recommended in Section 2.4, ensures that these power gains do not come at the cost of Type I error inflation.

The application of our framework to ROSMAP microglial snRNA-seq data demonstrates its practical value for AD research. Neither the frequentist-equivalent nor the non-informative Bayesian analysis identified statistically significant pathways, while both the informative and neuron-specific informative priors successfully recovered biologically meaningful signals. The gamma-secretase pathways identified under the informative prior are directly relevant to amyloid-beta production, a hallmark of AD pathology (Haapasalo and Kovacs, 2011; Dries and Yu, 2008), and the HP1 family pathway is consistent with recent evidence of chromatin remodeling dysregulation in AD brains (Neuner et al., 2019). Notably, the neuron-specific informative prior additionally identified the neurofilament axon growth pathway, a finding uniquely enabled by incorporating cell-type-specific prior knowledge, and consistent with the evidence of neurofilament dysregulation in AD pathology (Fernandez-Martos et al., 2015; Mitew et al., 2013). These results collectively demonstrate that AI-assisted prior elicitation can meaningfully increase the sensitivity of pathway analysis in snRNA-seq studies.

Although we illustrated the framework within the context of linear models and applied it only to the ROSMAP data for AD research in this paper, the framework is broadly applicable to a wide range of statistical models and genomic applications. To our knowledge, this represents the first systematic integration of large language model-based prior elicitation with the Bayesian-frequentist hybrid inference, offering a scalable and reproducible solution to one of the most persistent challenges in applied Bayesian analysis.

Looking forward, the proposed framework has significant potential to advance AD research and scRNA-seq data analysis. As the global burden of AD continues to grow with projections suggesting the number of affected individuals will double by 2060 (Fang et al., 2025), there is an unprecedented need for powerful and biologically informed analytical tools. Single-cell technologies continue to generate increasingly large and complex datasets from postmortem brain tissue across diverse brain regions and cell types (Mathys et al., 2024; Morabito et al., 2021), and the statistical challenge of detecting subtle transcriptional changes against this high-dimensional background remains difficult. The AI-assisted hybrid Bayesian framework directly addresses this challenge by incorporating the rich body of existing genetic, genomic, and proteomic evidence into the analysis, thereby amplifying the signal from genes with established biological relevance. As LLMs continue to improve in scientific reasoning, the quality and specificity of AI-elicited priors are expected to improve accordingly, making genome-wide Bayesian analysis with informative priors increasingly feasible across diverse omics platforms. Moreover, as more cell-type-specific transcriptomic datasets from diverse brain regions and AD stages become available, the framework can be readily applied to build a more comprehensive molecular map of AD progression, ultimately contributing to the identification of novel therapeutic targets and the development of precision medicine strategies for AD.

Despite these promising results, we identified three limitations of the proposed method, which point to future research directions: First, the AI-elicitation approach is subject to potential hallucination and biases inherent in the training corpus of the language model (Sun et al., 2023), and we recommend expert review of AI-generated priors for genes identified as statistically significant. Second, the framework currently requires an external dataset to estimate the prior variability, but such external data may not always be available. Finally, the current implementation uses a five-level ordinal scale to translate AI assessments into prior means. More flexible and continuous prior specification strategies are an important direction for future methodological development.

## A Appendix A

**Proofs**

### A.1 Proof of Theorem 1

To prove Theorem 1, it is sufficient to show that 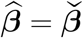, because for any 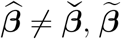 is between 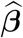 and 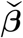 for all the three loss functions. With a uniform prior *π*(***β***) ∼ *U*[***a, b***], the posterior distribution of the full Bayesian model is

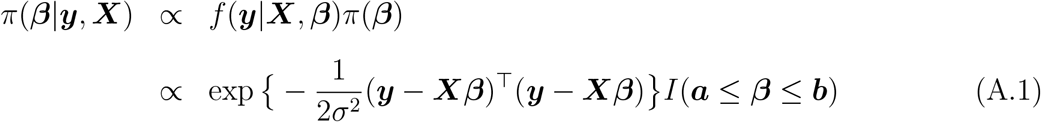

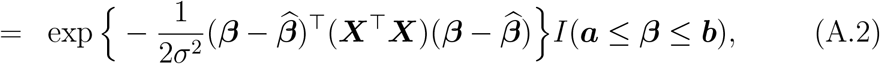

where 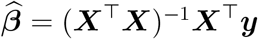. As a result, the posterior distribution of ***β*** is a truncated normal distribution on 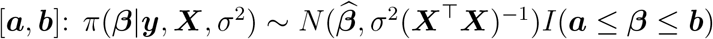.

Under the 0-1 loss, the Bayesian estimator is the posterior mode. Without loss of generality we assume ***β***^⋆^ ∈ [***a, b***]. Then using (A.1)

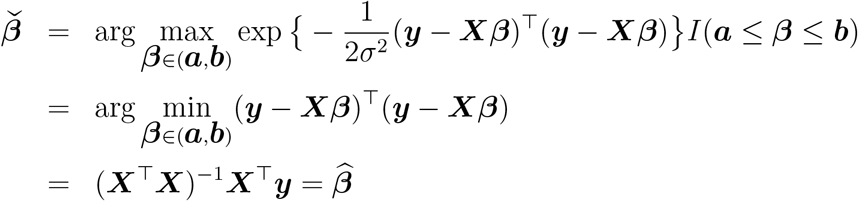

Under the absolute or squared error loss, the Bayesian estimator 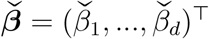 is the median or mean of the truncated posterior distribution in (A.2). So 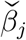 is the median or mean of the j-th marginal distribution of the truncated normal distribution 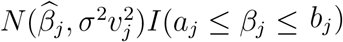, where 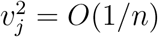 is the (*j, j*)-th element of (***X***^⊤^***X***)^−1^.

Recall that for a random variable *Z* ∼ *N*(*µ, σ*^2^)*I*(*a, b*), with −∞ ≤ *a* ≤ *Z* ≤ *b* ≤ ∞, then with 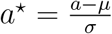 and 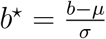

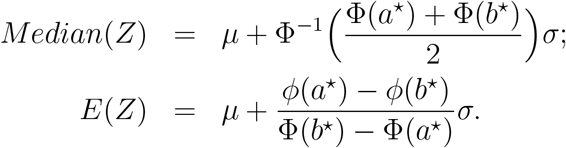

Using the above formula of *E*(*Z*), with the absolute error loss, the Bayes estimator is the posterior median 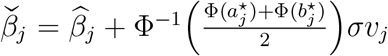, and with the squared error loss the Bayes estimator is the posterior mean 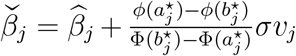, where 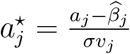 and 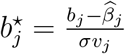 for all *j* ∈ {1, …, *d*}.

### A.2 Proof of Theorem 2

For the full Bayesian model, the posterior for ***β*** is

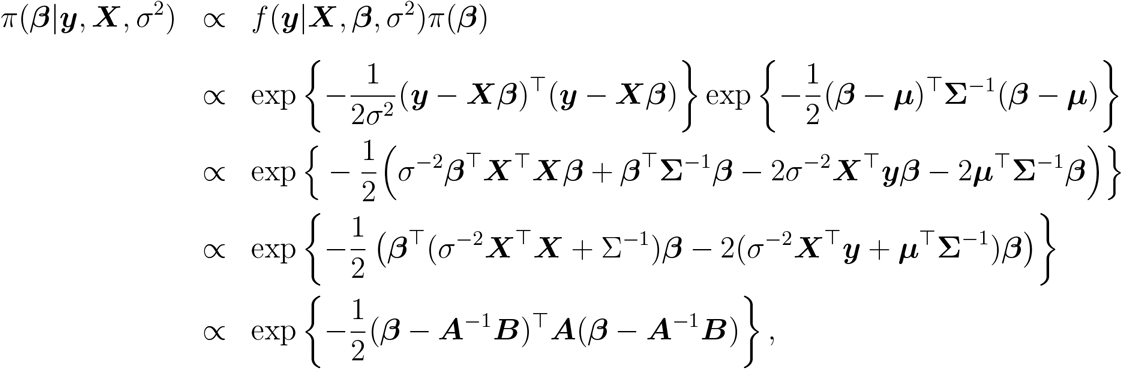

where ***A*** = (*σ*^−2^**X**^⊤^**X** + **Σ**^−1^), and ***B*** = *σ*^−2^**X**^⊤^**y** + ***µ***^⊤^**Σ**^−1^. as a result, *π*(***β***|***y, X***, *σ*^2^) ∼ *N*(***A***^−1^***B, A***^−1^). Because 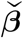 is the either posterior mean (with the squared error loss), or the posterior mode (with the 0-1 loss), or the posterior median (with the absolute error loss), the Bayesian inference point estimate 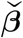 is the mean of the posterior distribution, i.e., 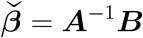.

For the hybrid model, denote ***X*** = (***X***_*B*_, ***X***_*F*_) and 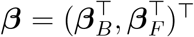. Conditioning on 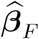 and *σ*^2^, the posterior for ***β***_*B*_ is

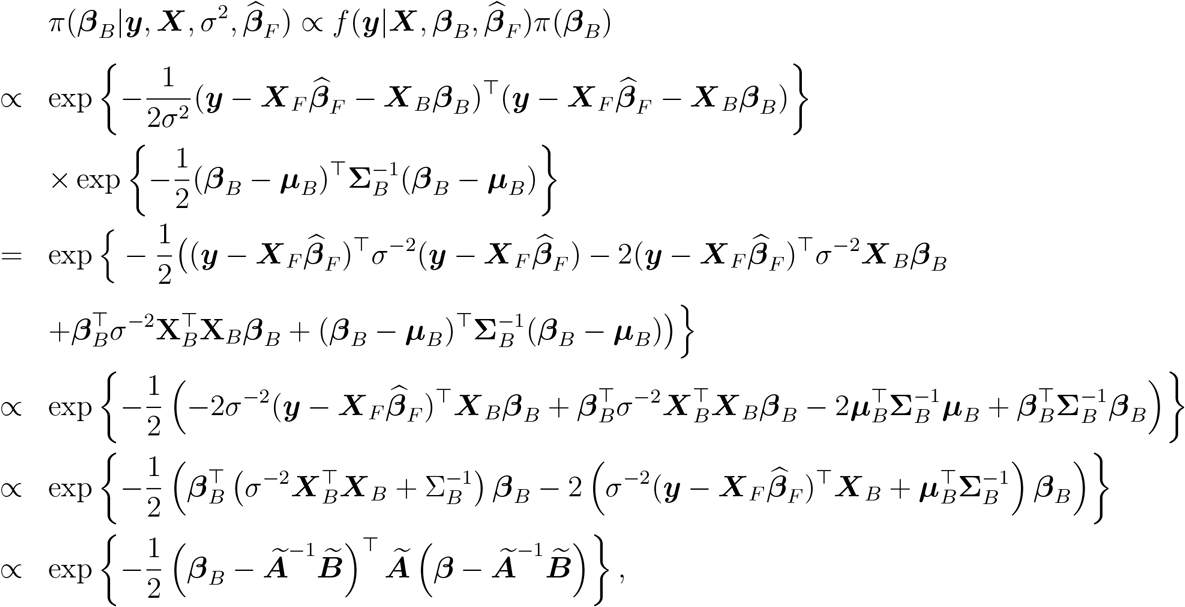

where 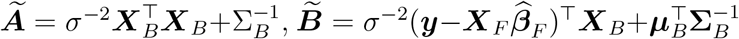. As a result, 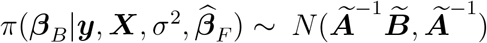 . Under the 0-1 loss, absolute error loss, and squared error loss, the hybrid estimate of ***β***_*B*_ is the posterior mode/median/mean of the above normal distribution, i.e., 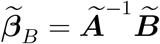. Given 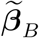 the hybrid estimate of the frequentist parameters is the least square estimate, i.e., 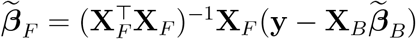.

### A.3 Proof of Corollary 1

Given that *π*(***β***_*F*_) is constant or non-informative and the prior *π*(***β***_*B*_) ∼ *N*(***µ***_*B*_, **Σ**_*B*_), we can define

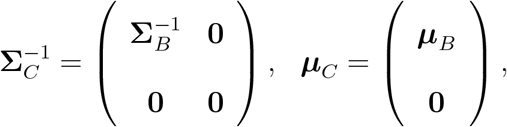

where 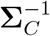 and ***µ***_*C*_ are of dimensions *d×d* and *d×*1, respectively. Then 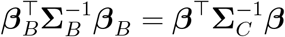 and 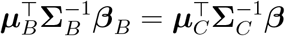.

The posterior distribution in the full Bayesian model can be derived as

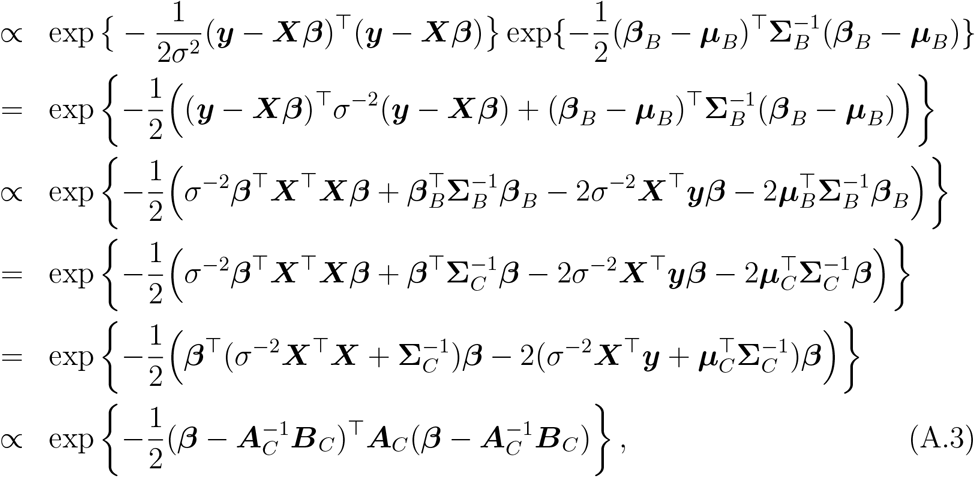

where 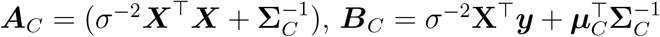. Given the density function form in (A.3), the posterior distribution of ***β*** is normal with mean 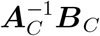 and variance 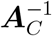, e.g., 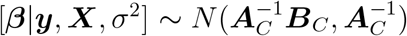, and the Bayes estimate of ***β*** under the squared error loss, 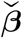, is the posterior mean, i.e., 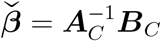.

In the hybrid inference, the posterior-likelihood can be written as

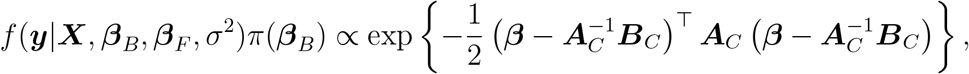

which is the same as (A.3). Under the squared error loss, the hybrid estimate 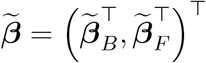 is obtained by the following mini-max procedure as in (1)

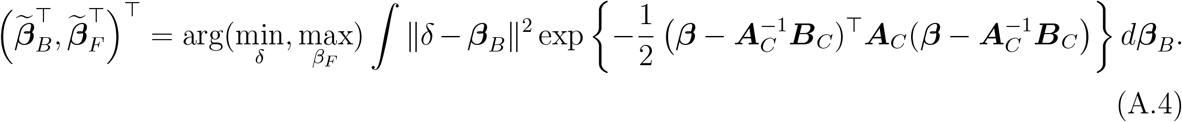

Take partial derivative with respect to *δ* on the above integral and set it to zero. Given a fixed ***β***_*F*_, 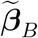 is the ***β***_*B*_ -marginal *mean* of the normal distribution in the above integrand (A.4), which is the same estimator as in the full Bayesian estimator with the same value of ***β***_*F*_ . Given 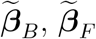 is chosen to minimize the objective function, i.e.,

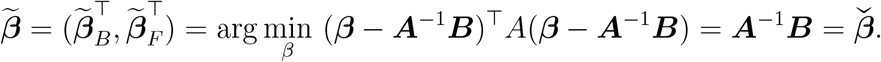

According to Yuan (2009), unconditional variances of the estimates from hybrid inference and Bayesian inference are asymptotically equivalent. ■

## B Appendix B

**The OpenAI API query in Python**

Below is the API prompt query regarding gene IFNB1’s general and neuron-specific informative priors for the association with AD:

~~~
import openai
client = openai.OpenAI()
response1 = client.chat.completions.create(
   model=“gpt-4o-2024-10-21”,
    messages=[
       {“role”: “user”, “content”: “Informative prior: Assess how strong “
       “the evidence of IFNB1 is associated with Alzheimer’s Disease? “
       “Please generate such evidence based on genetics, genomics, and “
       “proteomics. The evidence could be classified into level 1= none, “
       “level 2=weak, level 3=moderate, level 4=moderate to strong, and “
       “level 5=strong. Please strictly return 1, 2, 3, 4, or 5 based “
       “on your assessment.”}
    ],
    temperature=0.2,
    max_tokens=4000
)
response2 = client.chat.completions.create(
    model=“gpt-4o-2024-10-21”,
    messages=[
       {“role”: “user”, “content”: “Neuron-specific informative prior: “
       “Assess how strong the evidence of IFNB1 is associated with “
       “Alzheimer’s Disease in neurons? Please generate such evidence “
       “based on genetics, genomics, and proteomics. The evidence could “
       “be classified into level 1= none, level 2=weak, level 3=moderate, “
       “level 4=moderate to strong, and level 5=strong. Please strictly “
       “return 1, 2, 3, 4, or 5 based on your assessment.”}
    ],
    temperature=0.2,
    max_tokens=4000
)
~~~

## Acknowledgement

The data in the AD Knowledge Portal would not be possible without the participation of research volunteers and the contribution of data by collaborating researchers.

## Supplementary material

Supplementary materials are downloadable at the journal website.

## Data Availability

The snRNA-seq data are available on The Rush Alzheimer’s Disease Center Research Resource Sharing Hub at https://www.radc.rush.edu/docs/omics.htm or at Synapse (https://www.synapse.org/#!Synapse:syn18485175) under the doi 10.7303/syn18485175. The ROSMAP metadata can be accessed at https://www.synapse.org/#!Synapse:syn3157322. The results published here are in whole or in part based on data obtained from the AD Knowledge Portal (https://doi.org/10.7303/9618239).

