## Supplementary materials for "Generative AI-assisted Bayesian-frequentist Hybrid Inference in Single-cell RNA Sequencing Analysis for Genes Associated with Alzheimer’s Disease"

### Part 1. Additional pathway analysis results

| Pathway Analysis on Differentially Expressed Genes Using Non-informative prior | | | | | | | |
|---|---|---|---|---|---|---|---|
| | **r** | **R** | **n** | **N** | **Z** | **p-value** | **q-value** |
| Transcription_Role of heterochromatin protein 1 (HP1) family in transcriptional silencing | 4 | 76 | 40 | 12736 | 7.73 | 9.09e-05 | 0.12 |
| Neurophysiological process_Receptor-mediated axon growth repulsion | 4 | 76 | 46 | 12736 | 7.14 | 1.58E-04 | 0.12 |
| Pathway Analysis on Differentially Expressed Genes Using Informative prior | | | | | | | |
| | **r** | **R** | **n** | **N** | **Z-score** | **p-value** | **q-value** |
| Gamma-secretase proteolytic targets | 7 | 74 | 80 | 12736 | 9.64 | 3.79E-07 | 5.75E-04 |
| Gamma-Secretase regulation of neuronal cell development and function | 6 | 74 | 57 | 12736 | 9.90 | 8.99E-07 | 6.81E-04 |
| Neurophysiological process_ACM1, ACM3 and ACM5 signaling in the brain | 5 | 74 | 78 | 12736 | 6.79 | 8.77E-05 | 0.04 |
| Neurophysiological process Dynein-dynactin motor complex in axonal transport in neurons | 4 | 74 | 54 | 12736 | 6.61 | 2.67E-04 | 0.10 |
| Prolactin/ JAK2 signaling in breast cancer | 3 | 74 | 24 | 12736 | 7.69 | 3.49E-04 | 0.10 |
| Aberrant lipid trafficking and metabolism in age-related macular degeneration pathogenesis | 4 | 74 | 60 | 12736 | 6.22 | 4.00E-04 | 0.10 |
| Pathway Analysis on Differentially Expressed Genes Using Neuron-specific Informative prior | | | | | | | |
| | **r** | **R** | **n** | **N** | **Z** | **p-value** | **q-value** |
| Gamma-secretase proteolytic targets | 7 | 87 | 80 | 12736 | 8.79 | 1.15E-06 | 1.75E-03 |
| Gamma-Secretase regulation of neuronal cell development and function | 6 | 87 | 57 | 12736 | 9.04 | 2.34E-06 | 1.78E-03 |
| Transcription Role of heterochromatin protein 1 (HP1) family in transcriptional silencing | 5 | 87 | 40 | 12736 | 9.09 | 7.22E-06 | 3.65E-03 |
| Neurophysiological process_ACM1, ACM3 and ACM5 signaling in the brain | 5 | 87 | 78 | 12736 | 6.16 | 1.89E-04 | 0.07 |
| Cytoskeleton remodeling Neurofilaments in axon growth and synapses | 3 | 87 | 23 | 12736 | 7.20 | 4.94E-04 | 0.12 |
| Neurophysiological process Dynein-dynactin motor complex in axonal transport in neurons | 4 | 87 | 54 | 12736 | 6.01 | 4.95E-04 | 0.12 |
| Prolactin/ JAK2 signaling in breast cancer | 3 | 87 | 24 | 12736 | 7.03 | 5.62E-04 | 0.12 |
| Aberrant lipid trafficking and metabolism in age-related macular degeneration pathogenesis | 4 | 87 | 60 | 12736 | 5.64 | 7.40E-04 | 0.14 |

**r: intersection of ontology term with experiment list;**
**R: size of experiment list;**
**n: size of ontology term;**
**N: size of background list;**
**Z: z-score of enrichment;**
**p-value: hypergeometric test enrichment p-value;**
**q-value: FDR-adjusted p-value.**

### Part 2. Additional simulation results

Note: R programs for simulation are available to download at GitHub page: https://github.com/hangangtrue/AI_hybrid_inference

**Performance of $\beta_1$ Estimation in Scenarios A-F. True effect is the value of $\beta_1$ in the simulation.**

#### A: Weak data signal (SD(Y|X)=3), weak prior (SD($\beta_1$)=1), N=50

Sample size: n = 50; True effect = 0

$\beta_1$ Estimates (Mean (SD))

| Prior Effect | Frequentist | Bayesian | HB Conditional | HB Unconditional |
|---|---|---|---|---|
| 0.0 | -0.005 (0.888) | -0.002 (0.508) | -0.002 (0.508) | -0.002 (0.508) |
| 0.2 | 0.011 (0.881) | 0.092 (0.504) | 0.092 (0.504) | 0.092 (0.504) |
| 0.5 | -0.001 (0.889) | 0.214 (0.509) | 0.214 (0.509) | 0.214 (0.509) |
| 1.0 | -0.020 (0.879) | 0.419 (0.505) | 0.419 (0.505) | 0.419 (0.505) |

Standard Errors (Median [Q1, Q3])

| Prior Effect | Frequentist | Bayesian | HB Conditional | HB Unconditional |
|---|---|---|---|---|
| 0.0 | 0.870 [0.806, 0.933] | 0.656 [0.628, 0.682] | 0.423 [0.411, 0.433] | 0.641 [0.616, 0.664] |
| 0.2 | 0.870 [0.807, 0.936] | 0.656 [0.628, 0.683] | 0.423 [0.411, 0.434] | 0.641 [0.616, 0.664] |
| 0.5 | 0.869 [0.807, 0.933] | 0.656 [0.628, 0.682] | 0.423 [0.411, 0.433] | 0.640 [0.616, 0.663] |
| 1.0 | 0.872 [0.806, 0.938] | 0.657 [0.628, 0.684] | 0.423 [0.411, 0.434] | 0.642 [0.616, 0.665] |

Sample size: n = 50; True effect = 0.5

$\beta_1$ Estimates (Mean (SD))

| Prior Effect | Frequentist | Bayesian | HB Conditional | HB Unconditional |
|---|---|---|---|---|
| 0.0 | 0.492 (0.876) | 0.281 (0.501) | 0.281 (0.501) | 0.281 (0.501) |
| 0.2 | 0.501 (0.868) | 0.372 (0.495) | 0.372 (0.495) | 0.372 (0.495) |
| 0.5 | 0.509 (0.876) | 0.506 (0.500) | 0.506 (0.500) | 0.506 (0.500) |
| 1.0 | 0.497 (0.884) | 0.714 (0.507) | 0.713 (0.507) | 0.713 (0.507) |

Standard Errors (Median [Q1, Q3])

| Prior Effect | Frequentist | Bayesian | HB Conditional | HB Unconditional |
|---|---|---|---|---|
| 0.0 | 0.871 [0.807, 0.935] | 0.657 [0.628, 0.683] | 0.424 [0.411, 0.434] | 0.641 [0.616, 0.664] |
| 0.2 | 0.869 [0.806, 0.935] | 0.656 [0.627, 0.683] | 0.423 [0.411, 0.433] | 0.641 [0.615, 0.664] |
| 0.5 | 0.870 [0.807, 0.937] | 0.657 [0.628, 0.684] | 0.424 [0.411, 0.434] | 0.641 [0.616, 0.664] |
| 1.0 | 0.869 [0.804, 0.934] | 0.656 [0.627, 0.683] | 0.423 [0.411, 0.433] | 0.640 [0.615, 0.663] |

Sample size: n = 50; True effect = 1.0

$\beta_1$ Estimates (Mean (SD))

| Prior Effect | Frequentist | Bayesian | HB Conditional | HB Unconditional |
|---|---|---|---|---|
| 0.0 | 1.006 (0.874) | 0.573 (0.501) | 0.572 (0.501) | 0.572 (0.501) |
| 0.2 | 1.001 (0.873) | 0.657 (0.502) | 0.657 (0.502) | 0.657 (0.502) |
| 0.5 | 0.993 (0.877) | 0.781 (0.504) | 0.781 (0.504) | 0.781 (0.504) |
| 1.0 | 0.992 (0.873) | 0.995 (0.499) | 0.995 (0.499) | 0.995 (0.499) |

Standard Errors (Median [Q1, Q3])

| Prior Effect | Frequentist | Bayesian | HB Conditional | HB Unconditional |
|---|---|---|---|---|
| 0.0 | 0.870 [0.807, 0.936] | 0.656 [0.628, 0.683] | 0.424 [0.411, 0.434] | 0.641 [0.616, 0.664] |
| 0.2 | 0.870 [0.807, 0.933] | 0.656 [0.628, 0.682] | 0.423 [0.411, 0.433] | 0.640 [0.616, 0.664] |
| 0.5 | 0.868 [0.807, 0.934] | 0.656 [0.628, 0.683] | 0.423 [0.411, 0.433] | 0.640 [0.616, 0.663] |
| 1.0 | 0.870 [0.806, 0.934] | 0.656 [0.628, 0.683] | 0.423 [0.411, 0.433] | 0.641 [0.616, 0.664] |

### A: Weak data signal (SD(Y|X)=3), weak prior (SD($\beta_1$)=1), N=100

#### Sample size: n = 100; True effect = 0

##### $\beta_1$ Estimates (Mean (SD))

| Prior Effect | Frequentist | Bayesian | HB Conditional | HB Unconditional |
|---|---|---|---|---|
| 0.0 | 0.000 (0.607) | 0.000 (0.444) | 0.000 (0.444) | 0.000 (0.444) |
| 0.2 | 0.004 (0.608) | 0.057 (0.445) | 0.057 (0.445) | 0.057 (0.445) |
| 0.5 | 0.006 (0.610) | 0.139 (0.446) | 0.139 (0.446) | 0.139 (0.446) |
| 1.0 | 0.004 (0.617) | 0.272 (0.453) | 0.272 (0.453) | 0.272 (0.453) |

##### Standard Errors (Median [Q1, Q3])

| Prior Effect | Frequentist | Bayesian | HB Conditional | HB Unconditional |
|---|---|---|---|---|
| 0.0 | 0.608 [0.578, 0.637] | 0.519 [0.501, 0.537] | 0.354 [0.343, 0.365] | 0.515 [0.497, 0.533] |
| 0.2 | 0.608 [0.577, 0.638] | 0.519 [0.500, 0.538] | 0.355 [0.343, 0.366] | 0.515 [0.497, 0.533] |
| 0.5 | 0.606 [0.577, 0.637] | 0.518 [0.500, 0.537] | 0.354 [0.343, 0.365] | 0.514 [0.496, 0.533] |
| 1.0 | 0.607 [0.577, 0.637] | 0.519 [0.500, 0.538] | 0.354 [0.343, 0.366] | 0.515 [0.496, 0.533] |

#### Sample size: n = 100; True effect = 0.5

##### $\beta_1$ Estimates (Mean (SD))

| Prior Effect | Frequentist | Bayesian | HB Conditional | HB Unconditional |
|---|---|---|---|---|
| 0.0 | 0.492 (0.610) | 0.359 (0.446) | 0.359 (0.446) | 0.359 (0.446) |
| 0.2 | 0.491 (0.610) | 0.413 (0.446) | 0.413 (0.446) | 0.413 (0.446) |
| 0.5 | 0.494 (0.613) | 0.496 (0.447) | 0.496 (0.447) | 0.496 (0.447) |
| 1.0 | 0.513 (0.611) | 0.644 (0.447) | 0.644 (0.447) | 0.644 (0.447) |

##### Standard Errors (Median [Q1, Q3])

| Prior Effect | Frequentist | Bayesian | HB Conditional | HB Unconditional |
|---|---|---|---|---|
| 0.0 | 0.607 [0.577, 0.638] | 0.519 [0.500, 0.538] | 0.354 [0.342, 0.365] | 0.515 [0.497, 0.533] |
| 0.2 | 0.607 [0.577, 0.638] | 0.519 [0.500, 0.538] | 0.354 [0.342, 0.365] | 0.515 [0.496, 0.533] |
| 0.5 | 0.608 [0.578, 0.638] | 0.519 [0.501, 0.538] | 0.355 [0.343, 0.366] | 0.515 [0.497, 0.533] |
| 1.0 | 0.607 [0.577, 0.638] | 0.519 [0.500, 0.538] | 0.354 [0.343, 0.366] | 0.515 [0.497, 0.533] |

#### Sample size: n = 100; True effect = 1.0

##### $\beta_1$ Estimates (Mean (SD))

| Prior Effect | Frequentist | Bayesian | HB Conditional | HB Unconditional |
|---|---|---|---|---|
| 0.0 | 1.004 (0.606) | 0.734 (0.444) | 0.734 (0.444) | 0.734 (0.444) |
| 0.2 | 0.996 (0.608) | 0.781 (0.445) | 0.781 (0.445) | 0.781 (0.445) |
| 0.5 | 0.993 (0.601) | 0.860 (0.439) | 0.860 (0.439) | 0.860 (0.439) |
| 1.0 | 1.003 (0.611) | 1.002 (0.447) | 1.002 (0.447) | 1.002 (0.447) |

##### Standard Errors (Median [Q1, Q3]

| Prior Effect | Frequentist | Bayesian | HB Conditional | HB Unconditional |
|---|---|---|---|---|
| 0.0 | 0.606 [0.576, 0.638] | 0.518 [0.499, 0.538] | 0.354 [0.342, 0.365] | 0.515 [0.496, 0.533] |
| 0.2 | 0.607 [0.577, 0.638] | 0.519 [0.500, 0.538] | 0.354 [0.342, 0.366] | 0.515 [0.497, 0.533] |
| 0.5 | 0.607 [0.577, 0.638] | 0.519 [0.500, 0.538] | 0.354 [0.343, 0.366] | 0.515 [0.497, 0.533] |
| 1.0 | 0.607 [0.577, 0.637] | 0.519 [0.500, 0.537] | 0.354 [0.342, 0.365] | 0.515 [0.496, 0.533] |

### A: Weak data signal (SD(Y|X)=3), weak prior (SD($\beta_1$)=1), N=300

#### Sample size: n = 300; True effect = 0

$\beta_1$ Estimates (Mean (SD))

| Prior Effect | Frequentist | Bayesian | HB Conditional | HB Unconditional |
|---|---|---|---|---|
| 0.0 | 0.001 (0.346) | 0.001 (0.309) | 0.001 (0.309) | 0.001 (0.309) |
| 0.2 | 0.002 (0.350) | 0.024 (0.312) | 0.024 (0.312) | 0.024 (0.312) |
| 0.5 | 0.002 (0.353) | 0.056 (0.315) | 0.056 (0.315) | 0.056 (0.315) |
| 1.0 | 0.001 (0.347) | 0.109 (0.310) | 0.109 (0.310) | 0.109 (0.310) |

Standard Errors (Median [Q1, Q3])

| Prior Effect | Frequentist | Bayesian | HB Conditional | HB Unconditional |
|---|---|---|---|---|
| 0.0 | 0.348 [0.338, 0.358] | 0.329 [0.320, 0.337] | 0.230 [0.225, 0.236] | 0.328 [0.320, 0.336] |
| 0.2 | 0.348 [0.338, 0.358] | 0.329 [0.320, 0.337] | 0.231 [0.225, 0.236] | 0.328 [0.320, 0.336] |
| 0.5 | 0.348 [0.338, 0.358] | 0.328 [0.320, 0.337] | 0.230 [0.225, 0.236] | 0.328 [0.320, 0.336] |
| 1.0 | 0.347 [0.338, 0.357] | 0.328 [0.320, 0.337] | 0.230 [0.225, 0.236] | 0.328 [0.320, 0.336] |

#### Sample size: n = 300; True effect = 0.5

$\beta_1$ Estimates (Mean (SD))

| Prior Effect | Frequentist | Bayesian | HB Conditional | HB Unconditional |
|---|---|---|---|---|
| 0.0 | 0.504 (0.347) | 0.449 (0.310) | 0.449 (0.310) | 0.449 (0.310) |
| 0.2 | 0.498 (0.350) | 0.466 (0.312) | 0.466 (0.312) | 0.466 (0.312) |
| 0.5 | 0.499 (0.349) | 0.499 (0.311) | 0.499 (0.311) | 0.499 (0.311) |
| 1.0 | 0.500 (0.349) | 0.554 (0.311) | 0.554 (0.311) | 0.554 (0.311) |

Standard Errors (Median [Q1, Q3])

| Prior Effect | Frequentist | Bayesian | HB Conditional | HB Unconditional |
|---|---|---|---|---|
| 0.0 | 0.348 [0.338, 0.358] | 0.329 [0.320, 0.337] | 0.231 [0.225, 0.236] | 0.328 [0.320, 0.336] |
| 0.2 | 0.348 [0.338, 0.358] | 0.329 [0.320, 0.337] | 0.231 [0.225, 0.236] | 0.329 [0.320, 0.336] |
| 0.5 | 0.348 [0.338, 0.358] | 0.329 [0.320, 0.337] | 0.231 [0.225, 0.236] | 0.328 [0.320, 0.336] |
| 1.0 | 0.348 [0.338, 0.357] | 0.329 [0.320, 0.337] | 0.231 [0.225, 0.236] | 0.328 [0.320, 0.336] |

#### Sample size: n = 300; True effect = 1.0

$\beta_1$ Estimates (Mean (SD))

| Prior Effect | Frequentist | Bayesian | HB Conditional | HB Unconditional |
|---|---|---|---|---|
| 0.0 | 1.004 (0.351) | 0.896 (0.313) | 0.896 (0.313) | 0.896 (0.313) |
| 0.2 | 0.995 (0.350) | 0.909 (0.312) | 0.909 (0.312) | 0.909 (0.312) |
| 0.5 | 1.006 (0.347) | 0.951 (0.310) | 0.951 (0.310) | 0.951 (0.310) |
| 1.0 | 1.007 (0.353) | 1.006 (0.315) | 1.006 (0.315) | 1.006 (0.315) |

Standard Errors (Median [Q1, Q3])

| Prior Effect | Frequentist | Bayesian | HB Conditional | HB Unconditional |
|---|---|---|---|---|
| 0.0 | 0.348 [0.338, 0.358] | 0.328 [0.320, 0.337] | 0.230 [0.225, 0.236] | 0.328 [0.320, 0.336] |
| 0.2 | 0.348 [0.338, 0.358] | 0.329 [0.320, 0.337] | 0.231 [0.225, 0.236] | 0.328 [0.320, 0.336] |
| 0.5 | 0.347 [0.338, 0.357] | 0.328 [0.320, 0.336] | 0.230 [0.225, 0.236] | 0.328 [0.320, 0.336] |
| 1.0 | 0.348 [0.338, 0.358] | 0.329 [0.321, 0.337] | 0.231 [0.225, 0.236] | 0.328 [0.320, 0.337] |

### B: Strong data signal (SD(Y|X)=0.58), weak prior (SD($\beta_1$) =1), N=50

#### Sample size: n = 50; True effect = 0

##### $\beta_1$ Estimates (Mean (SD))

| Prior Effect | Frequentist | Bayesian | HB Conditional | HB Unconditional |
|---|---|---|---|---|
| 0.0 | 0.002 (0.171) | 0.002 (0.166) | 0.002 (0.166) | 0.002 (0.166) |
| 0.2 | 0.002 (0.169) | 0.007 (0.164) | 0.007 (0.164) | 0.007 (0.164) |
| 0.5 | -0.002 (0.170) | 0.012 (0.165) | 0.012 (0.165) | 0.012 (0.165) |
| 1.0 | 0.002 (0.170) | 0.030 (0.165) | 0.030 (0.165) | 0.030 (0.165) |

##### Standard Errors (Median [Q1, Q3])

| Prior Effect | Frequentist | Bayesian | HB Conditional | HB Unconditional |
|---|---|---|---|---|
| 0.0 | 0.168 [0.156, 0.181] | 0.166 [0.154, 0.178] | 0.114 [0.106, 0.122] | 0.166 [0.154, 0.178] |
| 0.2 | 0.168 [0.156, 0.181] | 0.166 [0.154, 0.178] | 0.113 [0.106, 0.122] | 0.166 [0.154, 0.178] |
| 0.5 | 0.167 [0.156, 0.180] | 0.165 [0.154, 0.178] | 0.113 [0.106, 0.121] | 0.165 [0.154, 0.178] |
| 1.0 | 0.168 [0.156, 0.181] | 0.166 [0.154, 0.178] | 0.114 [0.106, 0.122] | 0.166 [0.154, 0.178] |

#### Sample size: n = 50; True effect = 0.5

##### $\beta_1$ Estimates (Mean (SD))

| Prior Effect | Frequentist | Bayesian | HB Conditional | HB Unconditional |
|---|---|---|---|---|
| 0.0 | 0.499 (0.169) | 0.485 (0.164) | 0.485 (0.164) | 0.485 (0.164) |
| 0.2 | 0.499 (0.169) | 0.491 (0.164) | 0.491 (0.164) | 0.491 (0.164) |
| 0.5 | 0.502 (0.169) | 0.502 (0.164) | 0.502 (0.164) | 0.502 (0.164) |
| 1.0 | 0.502 (0.170) | 0.516 (0.165) | 0.516 (0.165) | 0.516 (0.165) |

##### Standard Errors (Median [Q1, Q3])

| Prior Effect | Frequentist | Bayesian | HB Conditional | HB Unconditional |
|---|---|---|---|---|
| 0.0 | 0.168 [0.156, 0.181] | 0.166 [0.154, 0.178] | 0.114 [0.106, 0.122] | 0.166 [0.154, 0.178] |
| 0.2 | 0.168 [0.156, 0.181] | 0.165 [0.154, 0.178] | 0.114 [0.106, 0.122] | 0.166 [0.154, 0.178] |
| 0.5 | 0.168 [0.156, 0.180] | 0.165 [0.154, 0.177] | 0.113 [0.106, 0.121] | 0.165 [0.154, 0.178] |
| 1.0 | 0.168 [0.155, 0.181] | 0.166 [0.154, 0.178] | 0.113 [0.106, 0.122] | 0.166 [0.154, 0.178] |

#### Sample size: n = 50; True effect = 1.0

##### $\beta_1$ Estimates (Mean (SD))

| Prior Effect | Frequentist | Bayesian | HB Conditional | HB Unconditional |
|---|---|---|---|---|
| 0.0 | 1.001 (0.167) | 0.973 (0.162) | 0.973 (0.162) | 0.973 (0.162) |
| 0.2 | 0.997 (0.168) | 0.975 (0.164) | 0.975 (0.164) | 0.975 (0.164) |
| 0.5 | 0.998 (0.170) | 0.984 (0.165) | 0.984 (0.165) | 0.984 (0.165) |
| 1.0 | 1.001 (0.171) | 1.001 (0.166) | 1.001 (0.166) | 1.001 (0.166) |

##### Standard Errors (Median [Q1, Q3])

| Prior Effect | Frequentist | Bayesian | HB Conditional | HB Unconditional |
|---|---|---|---|---|
| 0.0 | 0.168 [0.156, 0.181] | 0.166 [0.154, 0.178] | 0.114 [0.106, 0.122] | 0.166 [0.154, 0.178] |
| 0.2 | 0.168 [0.156, 0.181] | 0.166 [0.154, 0.178] | 0.114 [0.106, 0.122] | 0.166 [0.154, 0.178] |
| 0.5 | 0.169 [0.156, 0.182] | 0.166 [0.154, 0.179] | 0.114 [0.106, 0.122] | 0.166 [0.155, 0.179] |
| 1.0 | 0.168 [0.156, 0.181] | 0.166 [0.154, 0.178] | 0.114 [0.106, 0.122] | 0.166 [0.154, 0.178] |

### B: Strong data signal (SD(Y|X)=0.58), weak prior (SD($\beta_1$)=1), N=100

#### Sample size: n = 100; True effect = 0

$\beta_1$ Estimates (Mean (SD))

| Prior Effect | Frequentist | Bayesian | HB Conditional | HB Unconditional |
|---|---|---|---|---|
| 0.0 | 0.001 (0.119) | 0.001 (0.117) | 0.001 (0.117) | 0.001 (0.117) |
| 0.2 | 0.000 (0.118) | 0.003 (0.116) | 0.003 (0.116) | 0.003 (0.116) |
| 0.5 | 0.001 (0.119) | 0.008 (0.117) | 0.007 (0.117) | 0.007 (0.117) |
| 1.0 | 0.001 (0.118) | 0.014 (0.117) | 0.014 (0.117) | 0.014 (0.117) |

Standard Errors (Median [Q1, Q3])

| Prior Effect | Frequentist | Bayesian | HB Conditional | HB Unconditional |
|---|---|---|---|---|
| 0.0 | 0.117 [0.112, 0.123] | 0.117 [0.111, 0.122] | 0.081 [0.077, 0.085] | 0.117 [0.111, 0.122] |
| 0.2 | 0.117 [0.112, 0.123] | 0.117 [0.111, 0.122] | 0.081 [0.077, 0.085] | 0.117 [0.111, 0.122] |
| 0.5 | 0.118 [0.112, 0.123] | 0.117 [0.111, 0.122] | 0.081 [0.077, 0.085] | 0.117 [0.111, 0.122] |
| 1.0 | 0.117 [0.112, 0.123] | 0.116 [0.111, 0.122] | 0.081 [0.077, 0.085] | 0.116 [0.111, 0.122] |

#### Sample size: n = 100; True effect = 0.5

$\beta_1$ Estimates (Mean (SD))

| Prior Effect | Frequentist | Bayesian | HB Conditional | HB Unconditional |
|---|---|---|---|---|
| 0.0 | 0.501 (0.118) | 0.495 (0.116) | 0.495 (0.116) | 0.495 (0.116) |
| 0.2 | 0.498 (0.118) | 0.494 (0.116) | 0.494 (0.116) | 0.494 (0.116) |
| 0.5 | 0.502 (0.118) | 0.502 (0.117) | 0.502 (0.117) | 0.502 (0.117) |
| 1.0 | 0.498 (0.118) | 0.505 (0.116) | 0.505 (0.116) | 0.505 (0.116) |

Standard Errors (Median [Q1, Q3])

| Prior Effect | Frequentist | Bayesian | HB Conditional | HB Unconditional |
|---|---|---|---|---|
| 0.0 | 0.117 [0.111, 0.123] | 0.116 [0.111, 0.122] | 0.081 [0.077, 0.085] | 0.116 [0.111, 0.122] |
| 0.2 | 0.118 [0.112, 0.123] | 0.117 [0.111, 0.123] | 0.081 [0.077, 0.085] | 0.117 [0.111, 0.123] |
| 0.5 | 0.117 [0.111, 0.123] | 0.117 [0.111, 0.123] | 0.081 [0.077, 0.085] | 0.117 [0.111, 0.123] |
| 1.0 | 0.117 [0.112, 0.123] | 0.117 [0.111, 0.122] | 0.081 [0.077, 0.085] | 0.117 [0.111, 0.122] |

#### Sample size: n = 100; True effect = 1.0

$\beta_1$ Estimates (Mean (SD))

| Prior Effect | Frequentist | Bayesian | HB Conditional | HB Unconditional |
|---|---|---|---|---|
| 0.0 | 1.000 (0.118) | 0.986 (0.116) | 0.986 (0.116) | 0.986 (0.116) |
| 0.2 | 1.000 (0.118) | 0.989 (0.117) | 0.989 (0.117) | 0.989 (0.117) |
| 0.5 | 1.001 (0.118) | 0.994 (0.116) | 0.994 (0.116) | 0.994 (0.116) |
| 1.0 | 1.000 (0.117) | 1.000 (0.116) | 1.000 (0.116) | 1.000 (0.116) |

Standard Errors (Median [Q1, Q3])

| Prior Effect | Frequentist | Bayesian | HB Conditional | HB Unconditional |
|---|---|---|---|---|
| 0.0 | 0.118 [0.111, 0.123] | 0.117 [0.111, 0.122] | 0.081 [0.077, 0.085] | 0.117 [0.111, 0.122] |
| 0.2 | 0.117 [0.112, 0.124] | 0.117 [0.111, 0.123] | 0.081 [0.077, 0.085] | 0.117 [0.111, 0.123] |
| 0.5 | 0.117 [0.112, 0.123] | 0.117 [0.111, 0.122] | 0.081 [0.077, 0.085] | 0.117 [0.111, 0.122] |
| 1.0 | 0.117 [0.112, 0.123] | 0.117 [0.111, 0.122] | 0.081 [0.077, 0.085] | 0.117 [0.111, 0.122] |

### B: Strong data signal (SD(Y|X)=0.58), weak prior (SD($\beta_1$)=1), N=300

#### Sample size: n = 300; True effect = 0

$\beta_1$ Estimates (Mean (SD))

| Prior Effect | Frequentist | Bayesian | HB Conditional | HB Unconditional |
|---|---|---|---|---|
| 0.0 | -0.001 (0.067) | -0.001 (0.067) | -0.001 (0.067) | -0.001 (0.067) |
| 0.2 | -0.001 (0.067) | 0.000 (0.067) | 0.000 (0.067) | 0.000 (0.067) |
| 0.5 | 0.000 (0.068) | 0.002 (0.067) | 0.002 (0.067) | 0.002 (0.067) |
| 1.0 | -0.001 (0.067) | 0.004 (0.067) | 0.004 (0.067) | 0.004 (0.067) |

Standard Errors (Median [Q1, Q3])

| Prior Effect | Frequentist | Bayesian | HB Conditional | HB Unconditional |
|---|---|---|---|---|
| 0.0 | 0.067 [0.065, 0.069] | 0.067 [0.065, 0.069] | 0.047 [0.046, 0.048] | 0.067 [0.065, 0.069] |
| 0.2 | 0.067 [0.065, 0.069] | 0.067 [0.065, 0.069] | 0.047 [0.046, 0.049] | 0.067 [0.065, 0.069] |
| 0.5 | 0.067 [0.065, 0.069] | 0.067 [0.065, 0.069] | 0.047 [0.046, 0.048] | 0.067 [0.065, 0.069] |
| 1.0 | 0.067 [0.065, 0.069] | 0.067 [0.065, 0.069] | 0.047 [0.046, 0.049] | 0.067 [0.065, 0.069] |

#### Sample size: n = 300; True effect = 0.5

$\beta_1$ Estimates (Mean (SD))

| Prior Effect | Frequentist | Bayesian | HB Conditional | HB Unconditional |
|---|---|---|---|---|
| 0.0 | 0.499 (0.068) | 0.497 (0.068) | 0.497 (0.068) | 0.497 (0.068) |
| 0.2 | 0.500 (0.067) | 0.499 (0.067) | 0.499 (0.067) | 0.499 (0.067) |
| 0.5 | 0.499 (0.067) | 0.499 (0.067) | 0.499 (0.067) | 0.499 (0.067) |
| 1.0 | 0.499 (0.067) | 0.502 (0.067) | 0.502 (0.067) | 0.502 (0.067) |

Standard Errors (Median [Q1, Q3])

| Prior Effect | Frequentist | Bayesian | HB Conditional | HB Unconditional |
|---|---|---|---|---|
| 0.0 | 0.067 [0.065, 0.069] | 0.067 [0.065, 0.069] | 0.047 [0.046, 0.049] | 0.067 [0.065, 0.069] |
| 0.2 | 0.067 [0.065, 0.069] | 0.067 [0.065, 0.069] | 0.047 [0.046, 0.049] | 0.067 [0.065, 0.069] |
| 0.5 | 0.067 [0.065, 0.069] | 0.067 [0.065, 0.069] | 0.047 [0.046, 0.048] | 0.067 [0.065, 0.069] |
| 1.0 | 0.067 [0.065, 0.069] | 0.067 [0.065, 0.069] | 0.047 [0.046, 0.049] | 0.067 [0.065, 0.069] |

#### Sample size: n = 300; True effect = 1.0

$\beta_1$ Estimates (Mean (SD))

| Prior Effect | Frequentist | Bayesian | HB Conditional | HB Unconditional |
|---|---|---|---|---|
| 0.0 | 1.000 (0.067) | 0.995 (0.067) | 0.995 (0.067) | 0.995 (0.067) |
| 0.2 | 1.000 (0.067) | 0.996 (0.067) | 0.996 (0.067) | 0.996 (0.067) |
| 0.5 | 0.999 (0.067) | 0.997 (0.066) | 0.997 (0.066) | 0.997 (0.066) |
| 1.0 | 1.001 (0.068) | 1.001 (0.068) | 1.001 (0.068) | 1.001 (0.068) |

Standard Errors (Median [Q1, Q3])

| Prior Effect | Frequentist | Bayesian | HB Conditional | HB Unconditional |
|---|---|---|---|---|
| 0.0 | 0.067 [0.065, 0.069] | 0.067 [0.065, 0.069] | 0.047 [0.046, 0.048] | 0.067 [0.065, 0.069] |
| 0.2 | 0.067 [0.065, 0.069] | 0.067 [0.065, 0.069] | 0.047 [0.046, 0.049] | 0.067 [0.065, 0.069] |
| 0.5 | 0.067 [0.065, 0.069] | 0.067 [0.065, 0.069] | 0.047 [0.046, 0.048] | 0.067 [0.065, 0.069] |
| 1.0 | 0.067 [0.065, 0.069] | 0.067 [0.065, 0.069] | 0.047 [0.046, 0.049] | 0.067 [0.065, 0.069] |

### C: Weak data signal (SD(Y|X)=3), strong prior ($SD(\beta_1)=0.1$), N=50

#### Sample size: n = 50; True effect = 0

#### $\beta_1$ Estimates (Mean (SD))

| Prior Effect | Frequentist | Bayesian | HB Conditional | HB Unconditional |
|---|---|---|---|---|
| 0.0 | -0.009 (0.877) | -0.000 (0.012) | -0.000 (0.012) | -0.000 (0.012) |
| 0.2 | 0.006 (0.883) | 0.197 (0.012) | 0.197 (0.012) | 0.197 (0.012) |
| 0.5 | 0.001 (0.876) | 0.493 (0.012) | 0.493 (0.012) | 0.493 (0.012) |
| 1.0 | 0.007 (0.874) | 0.987 (0.012) | 0.987 (0.012) | 0.987 (0.012) |

#### Standard Errors (Median [Q1, Q3])

| Prior Effect | Frequentist | Bayesian | HB Conditional | HB Unconditional |
|---|---|---|---|---|
| 0.0 | 0.867 [0.805, 0.932] | 0.099 [0.099, 0.099] | 0.011 [0.011, 0.012] | 0.024 [0.022, 0.026] |
| 0.2 | 0.870 [0.806, 0.934] | 0.099 [0.099, 0.099] | 0.011 [0.011, 0.012] | 0.024 [0.022, 0.026] |
| 0.5 | 0.871 [0.808, 0.936] | 0.099 [0.099, 0.099] | 0.011 [0.011, 0.012] | 0.024 [0.022, 0.026] |
| 1.0 | 0.871 [0.807, 0.936] | 0.099 [0.099, 0.099] | 0.011 [0.011, 0.012] | 0.024 [0.022, 0.026] |

#### Sample size: n = 50; True effect = 0.5

#### $\beta_1$ Estimates (Mean (SD))

| Prior Effect | Frequentist | Bayesian | HB Conditional | HB Unconditional |
|---|---|---|---|---|
| 0.0 | 0.514 (0.864) | 0.007 (0.012) | 0.007 (0.012) | 0.007 (0.012) |
| 0.2 | 0.495 (0.885) | 0.204 (0.012) | 0.204 (0.012) | 0.204 (0.012) |
| 0.5 | 0.504 (0.878) | 0.500 (0.012) | 0.500 (0.012) | 0.500 (0.012) |
| 1.0 | 0.500 (0.878) | 0.993 (0.012) | 0.993 (0.012) | 0.993 (0.012) |

#### Standard Errors (Median [Q1, Q3])

| Prior Effect | Frequentist | Bayesian | HB Conditional | HB Unconditional |
|---|---|---|---|---|
| 0.0 | 0.869 [0.805, 0.935] | 0.099 [0.099, 0.099] | 0.011 [0.011, 0.012] | 0.024 [0.022, 0.026] |
| 0.2 | 0.870 [0.806, 0.936] | 0.099 [0.099, 0.099] | 0.011 [0.011, 0.012] | 0.024 [0.022, 0.026] |
| 0.5 | 0.868 [0.806, 0.934] | 0.099 [0.099, 0.099] | 0.011 [0.011, 0.012] | 0.024 [0.022, 0.026] |
| 1.0 | 0.870 [0.806, 0.934] | 0.099 [0.099, 0.099] | 0.011 [0.011, 0.012] | 0.024 [0.022, 0.026] |

#### Sample size: n = 50; True effect = 1.0

#### $\beta_1$ Estimates (Mean (SD))

| Prior Effect | Frequentist | Bayesian | HB Conditional | HB Unconditional |
|---|---|---|---|---|
| 0.0 | 1.010 (0.879) | 0.014 (0.013) | 0.014 (0.013) | 0.014 (0.013) |
| 0.2 | 1.012 (0.884) | 0.211 (0.012) | 0.211 (0.012) | 0.211 (0.012) |
| 0.5 | 1.003 (0.877) | 0.507 (0.012) | 0.507 (0.012) | 0.507 (0.012) |
| 1.0 | 0.992 (0.875) | 1.000 (0.012) | 1.000 (0.012) | 1.000 (0.012) |

#### Standard Errors (Median [Q1, Q3])

| Prior Effect | Frequentist | Bayesian | HB Conditional | HB Unconditional |
|---|---|---|---|---|
| 0.0 | 0.870 [0.807, 0.932] | 0.099 [0.099, 0.099] | 0.011 [0.011, 0.012] | 0.024 [0.022, 0.026] |
| 0.2 | 0.869 [0.805, 0.934] | 0.099 [0.099, 0.099] | 0.011 [0.011, 0.012] | 0.024 [0.022, 0.026] |
| 0.5 | 0.868 [0.807, 0.933] | 0.099 [0.099, 0.099] | 0.011 [0.011, 0.012] | 0.024 [0.022, 0.026] |
| 1.0 | 0.871 [0.809, 0.936] | 0.099 [0.099, 0.099] | 0.011 [0.011, 0.012] | 0.024 [0.022, 0.026] |

### C: Weak data signal (SD(Y|X)=3), strong prior (SD($\beta_1$)=0.1), N=100

#### Sample size: n = 100; True effect = 0

$\beta_1$ Estimates (Mean (SD))

| Prior Effect | Frequentist | Bayesian | HB Conditional | HB Unconditional |
|---|---|---|---|---|
| 0.0 | -0.008 (0.603) | -0.000 (0.016) | -0.000 (0.016) | -0.000 (0.016) |
| 0.2 | -0.004 (0.608) | 0.195 (0.016) | 0.195 (0.016) | 0.195 (0.016) |
| 0.5 | 0.006 (0.606) | 0.487 (0.017) | 0.487 (0.017) | 0.487 (0.017) |
| 1.0 | 0.006 (0.608) | 0.973 (0.017) | 0.973 (0.017) | 0.973 (0.017) |

Standard Errors (Median [Q1, Q3])

| Prior Effect | Frequentist | Bayesian | HB Conditional | HB Unconditional |
|---|---|---|---|---|
| 0.0 | 0.607 [0.578, 0.638] | 0.099 [0.099, 0.099] | 0.016 [0.015, 0.017] | 0.032 [0.031, 0.034] |
| 0.2 | 0.606 [0.577, 0.637] | 0.099 [0.099, 0.099] | 0.016 [0.015, 0.017] | 0.032 [0.031, 0.034] |
| 0.5 | 0.607 [0.576, 0.637] | 0.099 [0.099, 0.099] | 0.016 [0.015, 0.017] | 0.032 [0.031, 0.034] |
| 1.0 | 0.608 [0.577, 0.638] | 0.099 [0.099, 0.099] | 0.016 [0.015, 0.017] | 0.032 [0.031, 0.034] |

#### Sample size: n = 100; True effect = 0.5

$\beta_1$ Estimates (Mean (SD))

| Prior Effect | Frequentist | Bayesian | HB Conditional | HB Unconditional |
|---|---|---|---|---|
| 0.0 | 0.488 (0.611) | 0.013 (0.017) | 0.013 (0.017) | 0.013 (0.017) |
| 0.2 | 0.496 (0.617) | 0.208 (0.017) | 0.208 (0.017) | 0.208 (0.017) |
| 0.5 | 0.509 (0.611) | 0.500 (0.017) | 0.500 (0.017) | 0.500 (0.017) |
| 1.0 | 0.502 (0.608) | 0.987 (0.017) | 0.987 (0.017) | 0.987 (0.017) |

Standard Errors (Median [Q1, Q3])

| Prior Effect | Frequentist | Bayesian | HB Conditional | HB Unconditional |
|---|---|---|---|---|
| 0.0 | 0.607 [0.577, 0.637] | 0.099 [0.099, 0.099] | 0.016 [0.015, 0.017] | 0.032 [0.031, 0.034] |
| 0.2 | 0.607 [0.577, 0.638] | 0.099 [0.099, 0.099] | 0.016 [0.015, 0.017] | 0.032 [0.031, 0.034] |
| 0.5 | 0.607 [0.576, 0.637] | 0.099 [0.099, 0.099] | 0.016 [0.015, 0.017] | 0.032 [0.031, 0.034] |
| 1.0 | 0.606 [0.577, 0.637] | 0.099 [0.099, 0.099] | 0.016 [0.015, 0.017] | 0.032 [0.031, 0.034] |

#### Sample size: n = 100; True effect = 1.0

$\beta_1$ Estimates (Mean (SD))

| Prior Effect | Frequentist | Bayesian | HB Conditional | HB Unconditional |
|---|---|---|---|---|
| 0.0 | 1.004 (0.607) | 0.027 (0.017) | 0.027 (0.017) | 0.027 (0.017) |
| 0.2 | 0.998 (0.614) | 0.221 (0.017) | 0.221 (0.017) | 0.221 (0.017) |
| 0.5 | 0.991 (0.612) | 0.513 (0.017) | 0.513 (0.017) | 0.513 (0.017) |
| 1.0 | 0.996 (0.604) | 1.000 (0.016) | 1.000 (0.016) | 1.000 (0.016) |

Standard Errors (Median [Q1, Q3]

| Prior Effect | Frequentist | Bayesian | HB Conditional | HB Unconditional |
|---|---|---|---|---|
| 0.0 | 0.607 [0.578, 0.638] | 0.099 [0.099, 0.099] | 0.016 [0.015, 0.017] | 0.032 [0.031, 0.034] |
| 0.2 | 0.607 [0.576, 0.638] | 0.099 [0.099, 0.099] | 0.016 [0.015, 0.017] | 0.032 [0.031, 0.034] |
| 0.5 | 0.607 [0.578, 0.637] | 0.099 [0.099, 0.099] | 0.016 [0.015, 0.017] | 0.032 [0.031, 0.034] |
| 1.0 | 0.607 [0.577, 0.638] | 0.099 [0.099, 0.099] | 0.016 [0.015, 0.017] | 0.032 [0.031, 0.034] |

### C: Weak data signal (SD(Y|X)=3), strong prior (SD($\beta_1$)=0.1), N=300

#### Sample size: n = 300; True effect = 0

$\beta_1$ Estimates (Mean (SD))

| Prior Effect | Frequentist | Bayesian | HB Conditional | HB Unconditional |
|---|---|---|---|---|
| 0.0 | -0.002 (0.348) | -0.000 (0.027) | -0.000 (0.027) | -0.000 (0.027) |
| 0.2 | 0.000 (0.350) | 0.185 (0.027) | 0.185 (0.027) | 0.185 (0.027) |
| 0.5 | -0.003 (0.349) | 0.461 (0.027) | 0.461 (0.027) | 0.461 (0.027) |
| 1.0 | 0.003 (0.351) | 0.924 (0.028) | 0.924 (0.028) | 0.924 (0.028) |

Standard Errors (Median [Q1, Q3])

| Prior Effect | Frequentist | Bayesian | HB Conditional | HB Unconditional |
|---|---|---|---|---|
| 0.0 | 0.348 [0.338, 0.357] | 0.096 [0.096, 0.096] | 0.026 [0.026, 0.027] | 0.050 [0.049, 0.051] |
| 0.2 | 0.348 [0.338, 0.357] | 0.096 [0.096, 0.096] | 0.026 [0.026, 0.027] | 0.050 [0.049, 0.051] |
| 0.5 | 0.348 [0.338, 0.358] | 0.096 [0.096, 0.096] | 0.026 [0.026, 0.027] | 0.050 [0.049, 0.051] |
| 1.0 | 0.348 [0.338, 0.358] | 0.096 [0.096, 0.096] | 0.026 [0.026, 0.027] | 0.050 [0.049, 0.051] |

#### Sample size: n = 300; True effect = 0.5

$\beta_1$ Estimates (Mean (SD))

| Prior Effect | Frequentist | Bayesian | HB Conditional | HB Unconditional |
|---|---|---|---|---|
| 0.0 | 0.494 (0.341) | 0.038 (0.026) | 0.038 (0.026) | 0.038 (0.026) |
| 0.2 | 0.500 (0.346) | 0.223 (0.027) | 0.223 (0.027) | 0.223 (0.027) |
| 0.5 | 0.498 (0.349) | 0.500 (0.027) | 0.500 (0.027) | 0.500 (0.027) |
| 1.0 | 0.500 (0.350) | 0.962 (0.027) | 0.962 (0.027) | 0.962 (0.027) |

Standard Errors (Median [Q1, Q3])

| Prior Effect | Frequentist | Bayesian | HB Conditional | HB Unconditional |
|---|---|---|---|---|
| 0.0 | 0.348 [0.338, 0.358] | 0.096 [0.096, 0.096] | 0.026 [0.026, 0.027] | 0.050 [0.049, 0.051] |
| 0.2 | 0.348 [0.339, 0.358] | 0.096 [0.096, 0.096] | 0.026 [0.026, 0.027] | 0.050 [0.049, 0.051] |
| 0.5 | 0.348 [0.338, 0.358] | 0.096 [0.096, 0.096] | 0.026 [0.026, 0.027] | 0.050 [0.049, 0.051] |
| 1.0 | 0.348 [0.338, 0.357] | 0.096 [0.096, 0.096] | 0.026 [0.026, 0.027] | 0.050 [0.049, 0.051] |

#### Sample size: n = 300; True effect = 1.0

$\beta_1$ Estimates (Mean (SD))

| Prior Effect | Frequentist | Bayesian | HB Conditional | HB Unconditional |
|---|---|---|---|---|
| 0.0 | 0.997 (0.348) | 0.076 (0.027) | 0.076 (0.027) | 0.076 (0.027) |
| 0.2 | 1.004 (0.345) | 0.262 (0.027) | 0.262 (0.027) | 0.262 (0.027) |
| 0.5 | 0.996 (0.348) | 0.538 (0.027) | 0.538 (0.027) | 0.538 (0.027) |
| 1.0 | 0.999 (0.346) | 1.000 (0.027) | 1.000 (0.027) | 1.000 (0.027) |

Standard Errors (Median [Q1, Q3])

| Prior Effect | Frequentist | Bayesian | HB Conditional | HB Unconditional |
|---|---|---|---|---|
| 0.0 | 0.348 [0.338, 0.358] | 0.096 [0.096, 0.096] | 0.026 [0.026, 0.027] | 0.050 [0.049, 0.051] |
| 0.2 | 0.348 [0.338, 0.357] | 0.096 [0.096, 0.096] | 0.026 [0.026, 0.027] | 0.050 [0.049, 0.051] |
| 0.5 | 0.348 [0.338, 0.357] | 0.096 [0.096, 0.096] | 0.026 [0.026, 0.027] | 0.050 [0.049, 0.051] |
| 1.0 | 0.348 [0.338, 0.357] | 0.096 [0.096, 0.096] | 0.026 [0.026, 0.027] | 0.050 [0.049, 0.051] |

### D: Strong data signal (SD(Y|X)=0.58), strong prior (SD($\beta_1$)=0.1), N=50

#### Sample size: n = 50; True effect = 0

$\beta_1$ Estimates (Mean (SD))

| Prior Effect | Frequentist | Bayesian | HB Conditional | HB Unconditional |
|---|---|---|---|---|
| 0.0 | -0.000 (0.171) | -0.000 (0.046) | -0.000 (0.046) | -0.000 (0.046) |
| 0.2 | -0.000 (0.169) | 0.147 (0.046) | 0.147 (0.046) | 0.147 (0.046) |
| 0.5 | -0.002 (0.169) | 0.367 (0.050) | 0.367 (0.050) | 0.367 (0.050) |
| 1.0 | 0.004 (0.169) | 0.736 (0.063) | 0.736 (0.063) | 0.736 (0.063) |

Standard Errors (Median [Q1, Q3])

| Prior Effect | Frequentist | Bayesian | HB Conditional | HB Unconditional |
|---|---|---|---|---|
| 0.0 | 0.168 [0.156, 0.180] | 0.086 [0.084, 0.087] | 0.042 [0.041, 0.043] | 0.072 [0.071, 0.073] |
| 0.2 | 0.168 [0.156, 0.181] | 0.086 [0.084, 0.088] | 0.042 [0.041, 0.043] | 0.072 [0.071, 0.073] |
| 0.5 | 0.168 [0.156, 0.181] | 0.086 [0.084, 0.088] | 0.042 [0.041, 0.043] | 0.072 [0.071, 0.073] |
| 1.0 | 0.168 [0.156, 0.181] | 0.086 [0.084, 0.088] | 0.042 [0.041, 0.043] | 0.072 [0.071, 0.073] |

#### Sample size: n = 50; True effect = 0.5

$\beta_1$ Estimates (Mean (SD))

| Prior Effect | Frequentist | Bayesian | HB Conditional | HB Unconditional |
|---|---|---|---|---|
| 0.0 | 0.500 (0.171) | 0.132 (0.051) | 0.132 (0.051) | 0.132 (0.051) |
| 0.2 | 0.502 (0.171) | 0.280 (0.048) | 0.280 (0.048) | 0.280 (0.048) |
| 0.5 | 0.503 (0.170) | 0.501 (0.045) | 0.501 (0.045) | 0.501 (0.045) |
| 1.0 | 0.497 (0.169) | 0.867 (0.050) | 0.867 (0.050) | 0.867 (0.050) |

Standard Errors (Median [Q1, Q3])

| Prior Effect | Frequentist | Bayesian | HB Conditional | HB Unconditional |
|---|---|---|---|---|
| 0.0 | 0.168 [0.156, 0.180] | 0.086 [0.084, 0.087] | 0.042 [0.041, 0.043] | 0.072 [0.071, 0.073] |
| 0.2 | 0.168 [0.155, 0.180] | 0.086 [0.084, 0.087] | 0.042 [0.041, 0.043] | 0.072 [0.071, 0.073] |
| 0.5 | 0.168 [0.156, 0.181] | 0.086 [0.084, 0.088] | 0.042 [0.041, 0.043] | 0.072 [0.071, 0.073] |
| 1.0 | 0.169 [0.156, 0.181] | 0.086 [0.084, 0.088] | 0.042 [0.041, 0.043] | 0.072 [0.071, 0.073] |

#### Sample size: n = 50; True effect = 1.0

$\beta_1$ Estimates (Mean (SD))

| Prior Effect | Frequentist | Bayesian | HB Conditional | HB Unconditional |
|---|---|---|---|---|
| 0.0 | 0.997 (0.169) | 0.263 (0.062) | 0.263 (0.062) | 0.263 (0.062) |
| 0.2 | 1.000 (0.169) | 0.412 (0.057) | 0.412 (0.057) | 0.412 (0.057) |
| 0.5 | 1.002 (0.169) | 0.633 (0.050) | 0.633 (0.050) | 0.633 (0.050) |
| 1.0 | 1.004 (0.168) | 1.001 (0.045) | 1.001 (0.045) | 1.001 (0.045) |

Standard Errors (Median [Q1, Q3])

| Prior Effect | Frequentist | Bayesian | HB Conditional | HB Unconditional |
|---|---|---|---|---|
| 0.0 | 0.168 [0.156, 0.181] | 0.086 [0.084, 0.088] | 0.042 [0.041, 0.043] | 0.072 [0.071, 0.073] |
| 0.2 | 0.168 [0.156, 0.181] | 0.086 [0.084, 0.087] | 0.042 [0.041, 0.043] | 0.072 [0.071, 0.073] |
| 0.5 | 0.168 [0.156, 0.181] | 0.086 [0.084, 0.087] | 0.042 [0.041, 0.043] | 0.072 [0.071, 0.073] |
| 1.0 | 0.168 [0.156, 0.181] | 0.086 [0.084, 0.088] | 0.042 [0.041, 0.043] | 0.072 [0.071, 0.073] |

### D: Strong data signal (SD(Y|X)=0.58), strong prior (SD($\beta_1$)=0.1), N=100

#### Sample size: n = 100; True effect = 0

$\beta_1$ Estimates (Mean (SD))

| Prior Effect | Frequentist | Bayesian | HB Conditional | HB Unconditional |
|---|---|---|---|---|
| 0.0 | -0.003 (0.117) | -0.001 (0.050) | -0.001 (0.050) | -0.001 (0.050) |
| 0.2 | 0.002 (0.117) | 0.116 (0.050) | 0.116 (0.050) | 0.116 (0.050) |
| 0.5 | 0.000 (0.117) | 0.289 (0.053) | 0.289 (0.053) | 0.289 (0.053) |
| 1.0 | 0.000 (0.116) | 0.579 (0.061) | 0.579 (0.061) | 0.579 (0.061) |

Standard Errors (Median [Q1, Q3])

| Prior Effect | Frequentist | Bayesian | HB Conditional | HB Unconditional |
|---|---|---|---|---|
| 0.0 | 0.117 [0.112, 0.123] | 0.076 [0.074, 0.078] | 0.045 [0.045, 0.045] | 0.070 [0.069, 0.071] |
| 0.2 | 0.117 [0.112, 0.123] | 0.076 [0.075, 0.078] | 0.045 [0.045, 0.045] | 0.070 [0.069, 0.071] |
| 0.5 | 0.117 [0.111, 0.123] | 0.076 [0.074, 0.078] | 0.045 [0.045, 0.045] | 0.070 [0.069, 0.071] |
| 1.0 | 0.118 [0.112, 0.123] | 0.076 [0.075, 0.078] | 0.045 [0.045, 0.045] | 0.070 [0.069, 0.071] |

#### Sample size: n = 100; True effect = 0.5

$\beta_1$ Estimates (Mean (SD))

| Prior Effect | Frequentist | Bayesian | HB Conditional | HB Unconditional |
|---|---|---|---|---|
| 0.0 | 0.499 (0.119) | 0.210 (0.053) | 0.210 (0.053) | 0.210 (0.053) |
| 0.2 | 0.500 (0.117) | 0.326 (0.051) | 0.326 (0.051) | 0.326 (0.051) |
| 0.5 | 0.500 (0.117) | 0.500 (0.049) | 0.500 (0.049) | 0.500 (0.049) |
| 1.0 | 0.500 (0.117) | 0.790 (0.053) | 0.790 (0.053) | 0.790 (0.053) |

Standard Errors (Median [Q1, Q3])

| Prior Effect | Frequentist | Bayesian | HB Conditional | HB Unconditional |
|---|---|---|---|---|
| 0.0 | 0.117 [0.112, 0.123] | 0.076 [0.074, 0.078] | 0.045 [0.045, 0.045] | 0.070 [0.069, 0.071] |
| 0.2 | 0.118 [0.112, 0.123] | 0.076 [0.074, 0.078] | 0.045 [0.045, 0.045] | 0.070 [0.069, 0.071] |
| 0.5 | 0.117 [0.112, 0.123] | 0.076 [0.075, 0.078] | 0.045 [0.045, 0.045] | 0.070 [0.069, 0.071] |
| 1.0 | 0.117 [0.112, 0.123] | 0.076 [0.075, 0.078] | 0.045 [0.045, 0.045] | 0.070 [0.069, 0.071] |

#### Sample size: n = 100; True effect = 1.0

$\beta_1$ Estimates (Mean (SD))

| Prior Effect | Frequentist | Bayesian | HB Conditional | HB Unconditional |
|---|---|---|---|---|
| 0.0 | 1.002 (0.118) | 0.422 (0.061) | 0.422 (0.061) | 0.422 (0.061) |
| 0.2 | 0.999 (0.118) | 0.537 (0.058) | 0.537 (0.058) | 0.537 (0.058) |
| 0.5 | 1.002 (0.118) | 0.711 (0.053) | 0.711 (0.053) | 0.711 (0.053) |
| 1.0 | 1.000 (0.118) | 1.000 (0.050) | 1.000 (0.050) | 1.000 (0.050) |

Standard Errors (Median [Q1, Q3]

| Prior Effect | Frequentist | Bayesian | HB Conditional | HB Unconditional |
|---|---|---|---|---|
| 0.0 | 0.118 [0.112, 0.123] | 0.076 [0.075, 0.078] | 0.045 [0.045, 0.045] | 0.070 [0.069, 0.071] |
| 0.2 | 0.117 [0.112, 0.123] | 0.076 [0.074, 0.078] | 0.045 [0.045, 0.045] | 0.070 [0.069, 0.071] |
| 0.5 | 0.118 [0.112, 0.123] | 0.076 [0.074, 0.078] | 0.045 [0.045, 0.045] | 0.070 [0.069, 0.071] |
| 1.0 | 0.117 [0.112, 0.123] | 0.076 [0.075, 0.078] | 0.045 [0.045, 0.045] | 0.070 [0.069, 0.071] |

### D: Strong data signal (SD(Y|X)=0.58), strong prior (SD($\beta_1$)=0.1), N=300

#### Sample size: n = 300; True effect = 0

$\beta_1$ Estimates (Mean (SD))

| Prior Effect | Frequentist | Bayesian | HB Conditional | HB Unconditional |
|---|---|---|---|---|
| 0.0 | 0.000 (0.067) | 0.000 (0.046) | 0.000 (0.046) | 0.000 (0.046) |
| 0.2 | -0.001 (0.068) | 0.062 (0.047) | 0.062 (0.047) | 0.062 (0.047) |
| 0.5 | -0.001 (0.067) | 0.155 (0.047) | 0.155 (0.047) | 0.155 (0.047) |
| 1.0 | -0.000 (0.067) | 0.311 (0.050) | 0.311 (0.050) | 0.311 (0.050) |

Standard Errors (Median [Q1, Q3])

| Prior Effect | Frequentist | Bayesian | HB Conditional | HB Unconditional |
|---|---|---|---|---|
| 0.0 | 0.067 [0.065, 0.069] | 0.056 [0.055, 0.057] | 0.038 [0.037, 0.039] | 0.055 [0.054, 0.056] |
| 0.2 | 0.067 [0.065, 0.069] | 0.056 [0.055, 0.057] | 0.038 [0.037, 0.039] | 0.055 [0.054, 0.056] |
| 0.5 | 0.067 [0.065, 0.069] | 0.056 [0.055, 0.057] | 0.038 [0.037, 0.039] | 0.055 [0.054, 0.056] |
| 1.0 | 0.067 [0.065, 0.069] | 0.056 [0.055, 0.057] | 0.038 [0.037, 0.039] | 0.055 [0.054, 0.056] |

#### Sample size: n = 300; True effect = 0.5

$\beta_1$ Estimates (Mean (SD))

| Prior Effect | Frequentist | Bayesian | HB Conditional | HB Unconditional |
|---|---|---|---|---|
| 0.0 | 0.501 (0.067) | 0.345 (0.047) | 0.345 (0.047) | 0.345 (0.047) |
| 0.2 | 0.500 (0.067) | 0.406 (0.047) | 0.406 (0.047) | 0.406 (0.047) |
| 0.5 | 0.500 (0.066) | 0.500 (0.046) | 0.500 (0.046) | 0.500 (0.046) |
| 1.0 | 0.501 (0.068) | 0.656 (0.047) | 0.656 (0.047) | 0.656 (0.047) |

Standard Errors (Median [Q1, Q3])

| Prior Effect | Frequentist | Bayesian | HB Conditional | HB Unconditional |
|---|---|---|---|---|
| 0.0 | 0.067 [0.065, 0.069] | 0.056 [0.055, 0.057] | 0.038 [0.037, 0.039] | 0.055 [0.054, 0.056] |
| 0.2 | 0.067 [0.065, 0.069] | 0.056 [0.055, 0.057] | 0.038 [0.037, 0.039] | 0.055 [0.054, 0.056] |
| 0.5 | 0.067 [0.065, 0.069] | 0.056 [0.055, 0.057] | 0.038 [0.037, 0.039] | 0.055 [0.054, 0.056] |
| 1.0 | 0.067 [0.065, 0.069] | 0.056 [0.055, 0.057] | 0.038 [0.037, 0.039] | 0.055 [0.054, 0.056] |

#### Sample size: n = 300; True effect = 1.0

$\beta_1$ Estimates (Mean (SD))

| Prior Effect | Frequentist | Bayesian | HB Conditional | HB Unconditional |
|---|---|---|---|---|
| 0.0 | 1.001 (0.067) | 0.689 (0.049) | 0.689 (0.049) | 0.689 (0.049) |
| 0.2 | 1.000 (0.067) | 0.750 (0.048) | 0.750 (0.048) | 0.750 (0.048) |
| 0.5 | 0.999 (0.067) | 0.844 (0.047) | 0.844 (0.047) | 0.844 (0.047) |
| 1.0 | 1.000 (0.068) | 1.000 (0.047) | 1.000 (0.047) | 1.000 (0.047) |

Standard Errors (Median [Q1, Q3])

| Prior Effect | Frequentist | Bayesian | HB Conditional | HB Unconditional |
|---|---|---|---|---|
| 0.0 | 0.067 [0.065, 0.069] | 0.056 [0.055, 0.057] | 0.038 [0.037, 0.039] | 0.055 [0.054, 0.056] |
| 0.2 | 0.067 [0.065, 0.069] | 0.056 [0.055, 0.057] | 0.038 [0.037, 0.039] | 0.055 [0.054, 0.056] |
| 0.5 | 0.067 [0.065, 0.069] | 0.056 [0.055, 0.057] | 0.038 [0.037, 0.039] | 0.055 [0.054, 0.056] |
| 1.0 | 0.067 [0.065, 0.069] | 0.056 [0.055, 0.057] | 0.038 [0.037, 0.039] | 0.055 [0.054, 0.056] |

### E: Strong data signal (SD(Y|X)=0.58), Moderate prior (SD($\beta_1$)=0.7), N=50

#### Sample size: n = 50; True effect = 0

#### $\beta_1$ Estimates (Mean (SD))

| Prior Effect | Frequentist | Bayesian | HB Conditional | HB Unconditional |
|---|---|---|---|---|
| 0.0 | 0.002 (0.169) | 0.002 (0.160) | 0.002 (0.160) | 0.002 (0.160) |
| 0.2 | 0.001 (0.171) | 0.012 (0.161) | 0.012 (0.161) | 0.012 (0.161) |
| 0.5 | 0.001 (0.168) | 0.028 (0.158) | 0.028 (0.158) | 0.028 (0.158) |
| 1.0 | -0.000 (0.170) | 0.055 (0.161) | 0.055 (0.161) | 0.055 (0.161) |

#### Standard Errors (Median [Q1, Q3])

| Prior Effect | Frequentist | Bayesian | HB Conditional | HB Unconditional |
|---|---|---|---|---|
| 0.0 | 0.168 [0.156, 0.181] | 0.164 [0.152, 0.176] | 0.112 [0.105, 0.120] | 0.164 [0.153, 0.176] |
| 0.2 | 0.168 [0.156, 0.180] | 0.163 [0.152, 0.175] | 0.112 [0.105, 0.120] | 0.163 [0.152, 0.175] |
| 0.5 | 0.168 [0.156, 0.181] | 0.163 [0.152, 0.175] | 0.112 [0.105, 0.120] | 0.164 [0.152, 0.176] |
| 1.0 | 0.168 [0.156, 0.181] | 0.164 [0.152, 0.175] | 0.112 [0.105, 0.120] | 0.164 [0.152, 0.176] |

#### Sample size: n = 50; True effect = 0.5

#### $\beta_1$ Estimates (Mean (SD))

| Prior Effect | Frequentist | Bayesian | HB Conditional | HB Unconditional |
|---|---|---|---|---|
| 0.0 | 0.496 (0.170) | 0.468 (0.161) | 0.468 (0.161) | 0.468 (0.161) |
| 0.2 | 0.502 (0.168) | 0.485 (0.159) | 0.485 (0.159) | 0.485 (0.159) |
| 0.5 | 0.499 (0.170) | 0.499 (0.160) | 0.499 (0.160) | 0.499 (0.160) |
| 1.0 | 0.498 (0.170) | 0.525 (0.161) | 0.525 (0.161) | 0.525 (0.161) |

#### Standard Errors (Median [Q1, Q3])

| Prior Effect | Frequentist | Bayesian | HB Conditional | HB Unconditional |
|---|---|---|---|---|
| 0.0 | 0.168 [0.156, 0.181] | 0.164 [0.152, 0.175] | 0.112 [0.104, 0.120] | 0.164 [0.152, 0.175] |
| 0.2 | 0.168 [0.156, 0.181] | 0.164 [0.152, 0.175] | 0.112 [0.105, 0.120] | 0.164 [0.153, 0.175] |
| 0.5 | 0.168 [0.156, 0.181] | 0.163 [0.152, 0.175] | 0.112 [0.104, 0.120] | 0.164 [0.152, 0.175] |
| 1.0 | 0.168 [0.156, 0.181] | 0.163 [0.152, 0.175] | 0.112 [0.105, 0.120] | 0.164 [0.153, 0.175] |

#### Sample size: n = 50; True effect = 1.0

#### $\beta_1$ Estimates (Mean (SD))

| Prior Effect | Frequentist | Bayesian | HB Conditional | HB Unconditional |
|---|---|---|---|---|
| 0.0 | 1.001 (0.170) | 0.946 (0.161) | 0.946 (0.161) | 0.946 (0.161) |
| 0.2 | 0.999 (0.170) | 0.955 (0.161) | 0.955 (0.161) | 0.955 (0.161) |
| 0.5 | 1.001 (0.169) | 0.974 (0.160) | 0.974 (0.160) | 0.974 (0.160) |
| 1.0 | 1.002 (0.168) | 1.002 (0.158) | 1.002 (0.158) | 1.002 (0.158) |

#### Standard Errors (Median [Q1, Q3])

| Prior Effect | Frequentist | Bayesian | HB Conditional | HB Unconditional |
|---|---|---|---|---|
| 0.0 | 0.168 [0.156, 0.180] | 0.164 [0.152, 0.175] | 0.112 [0.104, 0.120] | 0.164 [0.152, 0.175] |
| 0.2 | 0.168 [0.155, 0.181] | 0.163 [0.152, 0.175] | 0.112 [0.104, 0.120] | 0.164 [0.152, 0.175] |
| 0.5 | 0.168 [0.156, 0.181] | 0.164 [0.152, 0.175] | 0.112 [0.105, 0.120] | 0.164 [0.152, 0.175] |
| 1.0 | 0.168 [0.156, 0.181] | 0.163 [0.152, 0.175] | 0.112 [0.105, 0.120] | 0.164 [0.152, 0.175] |

### E: Strong data signal (SD(Y|X)=0.58), Moderate prior (SD($\beta_1$)=0.7), N=100

#### Sample size: n = 100; True effect = 0

$\beta_1$ Estimates (Mean (SD))

| Prior Effect | Frequentist | Bayesian | HB Conditional | HB Unconditional |
|---|---|---|---|---|
| 0.0 | 0.002 (0.117) | 0.002 (0.114) | 0.002 (0.114) | 0.002 (0.114) |
| 0.2 | -0.001 (0.117) | 0.005 (0.114) | 0.005 (0.114) | 0.005 (0.114) |
| 0.5 | 0.002 (0.118) | 0.016 (0.114) | 0.016 (0.114) | 0.016 (0.114) |
| 1.0 | 0.001 (0.116) | 0.028 (0.113) | 0.028 (0.113) | 0.028 (0.113) |

Standard Errors (Median [Q1, Q3])

| Prior Effect | Frequentist | Bayesian | HB Conditional | HB Unconditional |
|---|---|---|---|---|
| 0.0 | 0.117 [0.111, 0.123] | 0.116 [0.110, 0.122] | 0.081 [0.077, 0.085] | 0.116 [0.110, 0.122] |
| 0.2 | 0.117 [0.112, 0.123] | 0.116 [0.110, 0.121] | 0.081 [0.077, 0.085] | 0.116 [0.110, 0.121] |
| 0.5 | 0.117 [0.112, 0.123] | 0.116 [0.110, 0.121] | 0.081 [0.077, 0.084] | 0.116 [0.110, 0.121] |
| 1.0 | 0.117 [0.112, 0.123] | 0.116 [0.110, 0.121] | 0.081 [0.077, 0.085] | 0.116 [0.110, 0.122] |

#### Sample size: n = 100; True effect = 0.5

$\beta_1$ Estimates (Mean (SD))

| Prior Effect | Frequentist | Bayesian | HB Conditional | HB Unconditional |
|---|---|---|---|---|
| 0.0 | 0.501 (0.119) | 0.487 (0.116) | 0.487 (0.116) | 0.487 (0.116) |
| 0.2 | 0.500 (0.116) | 0.491 (0.113) | 0.491 (0.113) | 0.491 (0.113) |
| 0.5 | 0.501 (0.118) | 0.501 (0.114) | 0.501 (0.114) | 0.501 (0.114) |
| 1.0 | 0.502 (0.117) | 0.515 (0.114) | 0.515 (0.114) | 0.515 (0.114) |

Standard Errors (Median [Q1, Q3])

| Prior Effect | Frequentist | Bayesian | HB Conditional | HB Unconditional |
|---|---|---|---|---|
| 0.0 | 0.117 [0.111, 0.123] | 0.116 [0.110, 0.121] | 0.081 [0.077, 0.084] | 0.116 [0.110, 0.121] |
| 0.2 | 0.117 [0.112, 0.123] | 0.116 [0.110, 0.121] | 0.081 [0.077, 0.084] | 0.116 [0.110, 0.121] |
| 0.5 | 0.117 [0.112, 0.123] | 0.116 [0.110, 0.121] | 0.081 [0.077, 0.084] | 0.116 [0.110, 0.121] |
| 1.0 | 0.117 [0.112, 0.123] | 0.116 [0.110, 0.121] | 0.081 [0.077, 0.084] | 0.116 [0.110, 0.121] |

#### Sample size: n = 100; True effect = 1.0

$\beta_1$ Estimates (Mean (SD))

| Prior Effect | Frequentist | Bayesian | HB Conditional | HB Unconditional |
|---|---|---|---|---|
| 0.0 | 1.002 (0.119) | 0.974 (0.115) | 0.974 (0.115) | 0.974 (0.115) |
| 0.2 | 0.999 (0.118) | 0.977 (0.115) | 0.977 (0.115) | 0.977 (0.115) |
| 0.5 | 1.002 (0.118) | 0.988 (0.114) | 0.988 (0.114) | 0.988 (0.114) |
| 1.0 | 1.000 (0.118) | 1.000 (0.115) | 1.000 (0.115) | 1.000 (0.115) |

Standard Errors (Median [Q1, Q3]

| Prior Effect | Frequentist | Bayesian | HB Conditional | HB Unconditional |
|---|---|---|---|---|
| 0.0 | 0.117 [0.111, 0.123] | 0.116 [0.110, 0.121] | 0.081 [0.077, 0.084] | 0.116 [0.110, 0.121] |
| 0.2 | 0.117 [0.112, 0.123] | 0.116 [0.110, 0.121] | 0.081 [0.077, 0.084] | 0.116 [0.110, 0.121] |
| 0.5 | 0.117 [0.112, 0.123] | 0.116 [0.110, 0.121] | 0.081 [0.077, 0.084] | 0.116 [0.110, 0.121] |
| 1.0 | 0.117 [0.111, 0.123] | 0.116 [0.110, 0.122] | 0.081 [0.077, 0.085] | 0.116 [0.110, 0.122] |

### E: Strong data signal (SD(Y|X)=0.58), Moderate prior (SD($\beta_1$)=0.7), N=300

#### Sample size: n = 300; True effect = 0

$\beta_1$ Estimates (Mean (SD))

| Prior Effect | Frequentist | Bayesian | HB Conditional | HB Unconditional |
|---|---|---|---|---|
| 0.0 | -0.000 (0.068) | -0.000 (0.067) | -0.000 (0.067) | -0.000 (0.067) |
| 0.2 | 0.002 (0.066) | 0.004 (0.066) | 0.004 (0.066) | 0.004 (0.066) |
| 0.5 | -0.000 (0.067) | 0.004 (0.066) | 0.004 (0.066) | 0.004 (0.066) |
| 1.0 | -0.001 (0.067) | 0.009 (0.067) | 0.009 (0.067) | 0.009 (0.067) |

Standard Errors (Median [Q1, Q3])

| Prior Effect | Frequentist | Bayesian | HB Conditional | HB Unconditional |
|---|---|---|---|---|
| 0.0 | 0.067 [0.065, 0.069] | 0.067 [0.065, 0.069] | 0.047 [0.046, 0.048] | 0.067 [0.065, 0.069] |
| 0.2 | 0.067 [0.065, 0.069] | 0.067 [0.065, 0.069] | 0.047 [0.046, 0.048] | 0.067 [0.065, 0.069] |
| 0.5 | 0.067 [0.065, 0.069] | 0.067 [0.065, 0.069] | 0.047 [0.046, 0.048] | 0.067 [0.065, 0.069] |
| 1.0 | 0.067 [0.065, 0.069] | 0.067 [0.065, 0.069] | 0.047 [0.046, 0.048] | 0.067 [0.065, 0.069] |

#### Sample size: n = 300; True effect = 0.5

$\beta_1$ Estimates (Mean (SD))

| Prior Effect | Frequentist | Bayesian | HB Conditional | HB Unconditional |
|---|---|---|---|---|
| 0.0 | 0.499 (0.067) | 0.494 (0.067) | 0.494 (0.067) | 0.494 (0.067) |
| 0.2 | 0.501 (0.068) | 0.498 (0.067) | 0.498 (0.067) | 0.498 (0.067) |
| 0.5 | 0.501 (0.068) | 0.501 (0.067) | 0.501 (0.067) | 0.501 (0.067) |
| 1.0 | 0.499 (0.067) | 0.504 (0.066) | 0.504 (0.066) | 0.504 (0.066) |

Standard Errors (Median [Q1, Q3])

| Prior Effect | Frequentist | Bayesian | HB Conditional | HB Unconditional |
|---|---|---|---|---|
| 0.0 | 0.067 [0.065, 0.069] | 0.067 [0.065, 0.069] | 0.047 [0.046, 0.048] | 0.067 [0.065, 0.069] |
| 0.2 | 0.067 [0.065, 0.069] | 0.067 [0.065, 0.069] | 0.047 [0.046, 0.048] | 0.067 [0.065, 0.069] |
| 0.5 | 0.067 [0.065, 0.069] | 0.067 [0.065, 0.069] | 0.047 [0.046, 0.048] | 0.067 [0.065, 0.069] |
| 1.0 | 0.067 [0.065, 0.069] | 0.067 [0.065, 0.069] | 0.047 [0.046, 0.048] | 0.067 [0.065, 0.069] |

#### Sample size: n = 300; True effect = 1.0

$\beta_1$ Estimates (Mean (SD))

| Prior Effect | Frequentist | Bayesian | HB Conditional | HB Unconditional |
|---|---|---|---|---|
| 0.0 | 1.000 (0.068) | 0.991 (0.067) | 0.991 (0.067) | 0.991 (0.067) |
| 0.2 | 1.000 (0.067) | 0.993 (0.066) | 0.993 (0.066) | 0.993 (0.066) |
| 0.5 | 1.000 (0.067) | 0.995 (0.067) | 0.995 (0.067) | 0.995 (0.067) |
| 1.0 | 1.000 (0.067) | 1.000 (0.066) | 1.000 (0.066) | 1.000 (0.066) |

Standard Errors (Median [Q1, Q3])

| Prior Effect | Frequentist | Bayesian | HB Conditional | HB Unconditional |
|---|---|---|---|---|
| 0.0 | 0.067 [0.065, 0.069] | 0.067 [0.065, 0.069] | 0.047 [0.046, 0.048] | 0.067 [0.065, 0.069] |
| 0.2 | 0.067 [0.065, 0.069] | 0.067 [0.065, 0.069] | 0.047 [0.046, 0.048] | 0.067 [0.065, 0.069] |
| 0.5 | 0.067 [0.065, 0.069] | 0.067 [0.065, 0.069] | 0.047 [0.046, 0.048] | 0.067 [0.065, 0.069] |
| 1.0 | 0.067 [0.065, 0.069] | 0.067 [0.065, 0.069] | 0.047 [0.046, 0.048] | 0.067 [0.065, 0.069] |

### F: Weak data signal (SD(Y|X)=3), Moderate prior (SD($\beta_1$)=0.7), N=50

#### Sample size: n = 50; True effect = 0

#### $\beta_1$ Estimates (Mean (SD))

| Prior Effect | Frequentist | Bayesian | HB Conditional | HB Unconditional |
|---|---|---|---|---|
| 0.0 | 0.005 (0.879) | 0.003 (0.351) | 0.003 (0.351) | 0.003 (0.351) |
| 0.2 | -0.002 (0.879) | 0.121 (0.350) | 0.121 (0.350) | 0.121 (0.350) |
| 0.5 | -0.002 (0.875) | 0.301 (0.351) | 0.301 (0.351) | 0.301 (0.351) |
| 1.0 | -0.005 (0.878) | 0.602 (0.353) | 0.602 (0.353) | 0.602 (0.353) |

#### Standard Errors (Median [Q1, Q3])

| Prior Effect | Frequentist | Bayesian | HB Conditional | HB Unconditional |
|---|---|---|---|---|
| 0.0 | 0.869 [0.806, 0.934] | 0.545 [0.529, 0.560] | 0.314 [0.312, 0.316] | 0.500 [0.492, 0.510] |
| 0.2 | 0.869 [0.805, 0.935] | 0.545 [0.528, 0.560] | 0.314 [0.312, 0.316] | 0.500 [0.492, 0.510] |
| 0.5 | 0.869 [0.806, 0.932] | 0.545 [0.529, 0.560] | 0.314 [0.312, 0.316] | 0.500 [0.492, 0.510] |
| 1.0 | 0.868 [0.806, 0.932] | 0.545 [0.528, 0.560] | 0.314 [0.312, 0.316] | 0.500 [0.492, 0.510] |

#### Sample size: n = 50; True effect = 0.5

#### $\beta_1$ Estimates (Mean (SD))

| Prior Effect | Frequentist | Bayesian | HB Conditional | HB Unconditional |
|---|---|---|---|---|
| 0.0 | 0.486 (0.873) | 0.193 (0.351) | 0.193 (0.351) | 0.193 (0.351) |
| 0.2 | 0.498 (0.882) | 0.319 (0.353) | 0.319 (0.353) | 0.319 (0.353) |
| 0.5 | 0.509 (0.879) | 0.504 (0.351) | 0.503 (0.351) | 0.503 (0.351) |
| 1.0 | 0.511 (0.880) | 0.807 (0.352) | 0.807 (0.352) | 0.807 (0.352) |

#### Standard Errors (Median [Q1, Q3])

| Prior Effect | Frequentist | Bayesian | HB Conditional | HB Unconditional |
|---|---|---|---|---|
| 0.0 | 0.869 [0.807, 0.931] | 0.545 [0.529, 0.559] | 0.314 [0.312, 0.316] | 0.500 [0.492, 0.510] |
| 0.2 | 0.869 [0.806, 0.935] | 0.545 [0.528, 0.560] | 0.314 [0.311, 0.316] | 0.500 [0.493, 0.510] |
| 0.5 | 0.868 [0.807, 0.933] | 0.545 [0.529, 0.560] | 0.314 [0.312, 0.316] | 0.500 [0.492, 0.510] |
| 1.0 | 0.867 [0.805, 0.934] | 0.545 [0.528, 0.560] | 0.314 [0.311, 0.316] | 0.500 [0.492, 0.510] |

#### Sample size: n = 50; True effect = 1.0

#### $\beta_1$ Estimates (Mean (SD))

| Prior Effect | Frequentist | Bayesian | HB Conditional | HB Unconditional |
|---|---|---|---|---|
| 0.0 | 0.997 (0.869) | 0.396 (0.350) | 0.395 (0.350) | 0.395 (0.350) |
| 0.2 | 1.015 (0.882) | 0.522 (0.353) | 0.522 (0.353) | 0.522 (0.353) |
| 0.5 | 1.004 (0.881) | 0.700 (0.352) | 0.700 (0.352) | 0.700 (0.352) |
| 1.0 | 0.990 (0.883) | 0.996 (0.353) | 0.996 (0.353) | 0.996 (0.353) |

#### Standard Errors (Median [Q1, Q3])

| Prior Effect | Frequentist | Bayesian | HB Conditional | HB Unconditional |
|---|---|---|---|---|
| 0.0 | 0.867 [0.805, 0.933] | 0.545 [0.528, 0.560] | 0.314 [0.312, 0.316] | 0.500 [0.492, 0.510] |
| 0.2 | 0.869 [0.807, 0.933] | 0.545 [0.529, 0.560] | 0.314 [0.312, 0.316] | 0.500 [0.493, 0.510] |
| 0.5 | 0.869 [0.805, 0.934] | 0.545 [0.528, 0.560] | 0.314 [0.312, 0.316] | 0.500 [0.492, 0.510] |
| 1.0 | 0.869 [0.804, 0.933] | 0.545 [0.528, 0.560] | 0.314 [0.312, 0.316] | 0.500 [0.492, 0.510] |

### F: Weak data signal (SD(Y|X)=3), Moderate prior (SD($\beta_1$)=0.7), N=100

#### Sample size: n = 100; True effect = 0

$\beta_1$ Estimates (Mean (SD))

| Prior Effect | Frequentist | Bayesian | HB Conditional | HB Unconditional |
|---|---|---|---|---|
| 0.0 | 0.001 (0.609) | -0.000 (0.349) | -0.000 (0.349) | -0.000 (0.349) |
| 0.2 | -0.001 (0.609) | 0.085 (0.349) | 0.085 (0.349) | 0.085 (0.349) |
| 0.5 | -0.004 (0.612) | 0.213 (0.350) | 0.213 (0.350) | 0.213 (0.350) |
| 1.0 | -0.003 (0.605) | 0.428 (0.347) | 0.428 (0.347) | 0.428 (0.347) |

Standard Errors (Median [Q1, Q3])

| Prior Effect | Frequentist | Bayesian | HB Conditional | HB Unconditional |
|---|---|---|---|---|
| 0.0 | 0.607 [0.577, 0.638] | 0.459 [0.445, 0.471] | 0.299 [0.293, 0.304] | 0.445 [0.433, 0.455] |
| 0.2 | 0.606 [0.577, 0.637] | 0.458 [0.445, 0.471] | 0.298 [0.293, 0.303] | 0.444 [0.433, 0.455] |
| 0.5 | 0.607 [0.577, 0.637] | 0.459 [0.445, 0.471] | 0.299 [0.293, 0.304] | 0.445 [0.434, 0.455] |
| 1.0 | 0.608 [0.578, 0.638] | 0.459 [0.446, 0.471] | 0.299 [0.293, 0.304] | 0.445 [0.434, 0.455] |

#### Sample size: n = 100; True effect = 0.5

$\beta_1$ Estimates (Mean (SD))

| Prior Effect | Frequentist | Bayesian | HB Conditional | HB Unconditional |
|---|---|---|---|---|
| 0.0 | 0.504 (0.614) | 0.288 (0.351) | 0.288 (0.351) | 0.288 (0.351) |
| 0.2 | 0.511 (0.610) | 0.378 (0.349) | 0.378 (0.349) | 0.378 (0.349) |
| 0.5 | 0.499 (0.607) | 0.499 (0.347) | 0.499 (0.347) | 0.499 (0.347) |
| 1.0 | 0.493 (0.614) | 0.710 (0.352) | 0.710 (0.352) | 0.710 (0.352) |

Standard Errors (Median [Q1, Q3])

| Prior Effect | Frequentist | Bayesian | HB Conditional | HB Unconditional |
|---|---|---|---|---|
| 0.0 | 0.608 [0.578, 0.638] | 0.459 [0.446, 0.472] | 0.299 [0.293, 0.304] | 0.445 [0.434, 0.455] |
| 0.2 | 0.607 [0.577, 0.638] | 0.459 [0.445, 0.471] | 0.299 [0.293, 0.303] | 0.445 [0.433, 0.455] |
| 0.5 | 0.607 [0.577, 0.637] | 0.458 [0.445, 0.471] | 0.298 [0.293, 0.303] | 0.445 [0.434, 0.455] |
| 1.0 | 0.607 [0.577, 0.638] | 0.459 [0.445, 0.471] | 0.299 [0.293, 0.304] | 0.445 [0.434, 0.455] |

#### Sample size: n = 100; True effect = 1.0

$\beta_1$ Estimates (Mean (SD))

| Prior Effect | Frequentist | Bayesian | HB Conditional | HB Unconditional |
|---|---|---|---|---|
| 0.0 | 0.999 (0.607) | 0.571 (0.349) | 0.570 (0.349) | 0.570 (0.349) |
| 0.2 | 1.000 (0.605) | 0.657 (0.347) | 0.657 (0.347) | 0.657 (0.347) |
| 0.5 | 0.993 (0.610) | 0.782 (0.350) | 0.782 (0.350) | 0.782 (0.350) |
| 1.0 | 1.006 (0.609) | 1.004 (0.349) | 1.003 (0.349) | 1.003 (0.349) |

Standard Errors (Median [Q1, Q3]

| Prior Effect | Frequentist | Bayesian | HB Conditional | HB Unconditional |
|---|---|---|---|---|
| 0.0 | 0.607 [0.577, 0.639] | 0.459 [0.445, 0.472] | 0.299 [0.293, 0.304] | 0.445 [0.433, 0.455] |
| 0.2 | 0.607 [0.577, 0.637] | 0.458 [0.445, 0.471] | 0.299 [0.293, 0.304] | 0.444 [0.434, 0.455] |
| 0.5 | 0.607 [0.577, 0.636] | 0.458 [0.445, 0.471] | 0.299 [0.293, 0.303] | 0.445 [0.433, 0.455] |
| 1.0 | 0.607 [0.577, 0.637] | 0.459 [0.445, 0.471] | 0.299 [0.293, 0.303] | 0.444 [0.434, 0.455] |

### F: Weak data signal (SD(Y|X)=3), Moderate prior (SD($\beta_1$)=0.7), N=300

#### Sample size: n = 300; True effect = 0

$\beta_1$ Estimates (Mean (SD))

| Prior Effect | Frequentist | Bayesian | HB Conditional | HB Unconditional |
|---|---|---|---|---|
| 0.0 | 0.006 (0.349) | 0.005 (0.280) | 0.005 (0.280) | 0.005 (0.280) |
| 0.2 | 0.002 (0.348) | 0.041 (0.279) | 0.041 (0.279) | 0.041 (0.279) |
| 0.5 | -0.004 (0.352) | 0.096 (0.282) | 0.096 (0.282) | 0.096 (0.282) |
| 1.0 | -0.001 (0.348) | 0.197 (0.279) | 0.197 (0.279) | 0.197 (0.279) |

Standard Errors (Median [Q1, Q3])

| Prior Effect | Frequentist | Bayesian | HB Conditional | HB Unconditional |
|---|---|---|---|---|
| 0.0 | 0.348 [0.338, 0.358] | 0.311 [0.304, 0.318] | 0.217 [0.212, 0.221] | 0.310 [0.303, 0.317] |
| 0.2 | 0.348 [0.338, 0.357] | 0.312 [0.304, 0.318] | 0.217 [0.212, 0.221] | 0.310 [0.303, 0.317] |
| 0.5 | 0.348 [0.338, 0.358] | 0.311 [0.304, 0.318] | 0.217 [0.212, 0.221] | 0.310 [0.303, 0.317] |
| 1.0 | 0.348 [0.338, 0.358] | 0.311 [0.304, 0.318] | 0.217 [0.212, 0.221] | 0.310 [0.303, 0.317] |

#### Sample size: n = 300; True effect = 0.5

$\beta_1$ Estimates (Mean (SD))

| Prior Effect | Frequentist | Bayesian | HB Conditional | HB Unconditional |
|---|---|---|---|---|
| 0.0 | 0.502 (0.349) | 0.403 (0.280) | 0.403 (0.280) | 0.403 (0.280) |
| 0.2 | 0.494 (0.350) | 0.436 (0.280) | 0.436 (0.280) | 0.436 (0.280) |
| 0.5 | 0.497 (0.348) | 0.498 (0.279) | 0.498 (0.279) | 0.498 (0.279) |
| 1.0 | 0.500 (0.343) | 0.599 (0.276) | 0.599 (0.276) | 0.599 (0.276) |

Standard Errors (Median [Q1, Q3])

| Prior Effect | Frequentist | Bayesian | HB Conditional | HB Unconditional |
|---|---|---|---|---|
| 0.0 | 0.348 [0.338, 0.357] | 0.311 [0.304, 0.318] | 0.217 [0.212, 0.221] | 0.310 [0.303, 0.316] |
| 0.2 | 0.348 [0.338, 0.358] | 0.311 [0.304, 0.318] | 0.217 [0.212, 0.221] | 0.310 [0.303, 0.317] |
| 0.5 | 0.348 [0.339, 0.358] | 0.312 [0.305, 0.319] | 0.217 [0.212, 0.221] | 0.310 [0.303, 0.317] |
| 1.0 | 0.348 [0.338, 0.357] | 0.311 [0.304, 0.318] | 0.217 [0.212, 0.221] | 0.310 [0.303, 0.317] |

#### Sample size: n = 300; True effect = 1.0

$\beta_1$ Estimates (Mean (SD))

| Prior Effect | Frequentist | Bayesian | HB Conditional | HB Unconditional |
|---|---|---|---|---|
| 0.0 | 1.002 (0.350) | 0.803 (0.281) | 0.803 (0.281) | 0.803 (0.281) |
| 0.2 | 1.004 (0.350) | 0.844 (0.281) | 0.844 (0.281) | 0.844 (0.281) |
| 0.5 | 1.001 (0.349) | 0.902 (0.280) | 0.902 (0.280) | 0.902 (0.280) |
| 1.0 | 0.997 (0.346) | 0.997 (0.278) | 0.997 (0.278) | 0.997 (0.278) |

Standard Errors (Median [Q1, Q3])

| Prior Effect | Frequentist | Bayesian | HB Conditional | HB Unconditional |
|---|---|---|---|---|
| 0.0 | 0.348 [0.338, 0.357] | 0.311 [0.305, 0.318] | 0.217 [0.212, 0.221] | 0.310 [0.303, 0.317] |
| 0.2 | 0.348 [0.338, 0.358] | 0.312 [0.305, 0.319] | 0.217 [0.212, 0.221] | 0.310 [0.303, 0.317] |
| 0.5 | 0.348 [0.338, 0.358] | 0.311 [0.304, 0.318] | 0.217 [0.212, 0.221] | 0.310 [0.303, 0.317] |
| 1.0 | 0.348 [0.338, 0.358] | 0.312 [0.304, 0.319] | 0.217 [0.212, 0.221] | 0.310 [0.303, 0.317] |
